# Oral live attenuated polio vaccines induce enhanced T-cell responses with broad antigen recognition compared to inactivated polio vaccines

**DOI:** 10.1101/2025.05.20.25328004

**Authors:** Julia Snyder, Marni Slavik, Patrick Harvey, Roxana Del Rio-Guerra, Bernardo A. Mainou, Alessandro Sette, Beth D. Kirkpatrick, Alba Grifoni, Jessica W. Crothers

## Abstract

Little is known about the immunologic mechanisms responsible for observed differences in mucosal immunity following vaccination with oral live attenuated polio vaccines (OPVs) compared to inactivated polio vaccines (IPVs). Here, we used a flow cytometric activation-induced marker (AIM)-based approach to investigate vaccine-related differences in T cell response using peripheral blood samples from healthy adults enrolled in two polio vaccine trials. Our findings indicate that vaccination with OPVs (1) enhances CD4+ T cell responses, and (2) expands CD4+ and CD8+ antigen recognition of non-structural proteins. Fecal shedding of OPVs was associated with enhanced T cell responses, and primary CD8+ T cell responses appear to correlate with early control of fecal viral shedding. Our results indicate that OPVs induce broad cellular immunity to polioviruses, which may help explain their enhanced ability to stimulate effective pathogen-specific mucosal immunity.

## Introduction

Poliomyelitis is caused by three serotypes (1,2,3) of poliovirus (PV), which caused >400,000 cases of paralytic disease per year before the introduction of vaccines in the 1950s(1). With the success of trivalent oral and inactivated vaccine formulations, rates of paralytic disease fell precipitously and, along with declaration of the WHO Global Polio Eradication Initiative in 1988, there was much anticipation that polio would be the 2^nd^ human disease to be fully eradiated in the early 21^st^ century. However, despite herculean efforts, new and evolving challenges in the epidemiology of polio have arisen. Continued use of live attenuated oral polio vaccines (OPVs), which can revert to neurovirulence, combined with ongoing fecal shedding of poliovirus strains in un- or under-vaccinated populations have resulted in continued viral transmission and maintenance of environmental reservoirs of both wild-type and vaccine derived viral strains.

While both oral live attenuated (Sabin or novel, OPV) and inactivated (Sabin or Salk, IPV) polio vaccine formulations induce robust neutralizing antibody responses and protect from symptomatic disease, including paralysis (2–4), only OPVs stimulate sufficient immunity at mucosal sites to reduce fecal shedding of wild-type or vaccine viruses and limit viral transmission. Continued OPV replication leads to accumulation of viral mutations within the genetic regions of attenuation and subsequent circulation of neurovirulent vaccine-derived viral strains, which now account for >99.9% of paralytic cases of polio cases worldwide(5) (6–8). The paradoxical rise in the global incidence of paralytic poliomyelitis caused by vaccine derived OPV2 strains (vdPV2) following removal of OPV2 from trivalent formulations in 2016(2, 9, 10), emphasized the importance of OPV-induced control of viral replication (herein used as a proxy measure of effective mucosal immunity) and reduction of ongoing disease transmission across populations.

The immunological mechanisms responsible for differences in OPV versus IPV-induced mucosal immunity underpinning control of PV fecal shedding, however, are poorly understood. Unlike IPVs, OPVs stimulate immune responses through direct infection of mucosal tissues. Like wild-type PV strains, OPVs initiate cellular infection via binding to the poliovirus receptor (PVR, CD155), a cell adhesion molecule (CD155) highly expressed along the epithelial surfaces of the human alimentary tract and on key antigen presenting cells (APCs), including microfold (M cells) and dendritic cells (DCs), which are both permissive to PV infection(11, 12). Infection activates the innate immune system, providing early protection and initial engagement with the adaptive immune system. Robust serotype-specific serum neutralizing antibody (SNA) responses are detected 2 to 4 weeks following infection with either wildtype or OPV viral strains. Mucosal antibody responses (fecal Immunoglobulin, IgA and IgG) are detected in the stools of OPV-vaccinated infants, but only weakly predict the likelihood or amount of viral shedding upon subsequent OPV challenge(13, 14). In addition, although robust humoral responses are observed across age ranges, recent studies reveal startling age-related differences in fecal mucosal antibody production and despite high levels of fecal shedding, adults fail to mount fecal antibody responses following initial OPV exposure(15). Taken together, these data suggest that immunologic mechanisms other than humoral antibody responses likely play a role in generation of effective mucosal immunity to PV(16).

The role of T cells in antiviral immunity to PVs is underexplored in human populations despite evidence in animal models suggesting that CD4+ T cells are important for the generation of coordinated, effective memory responses to PV at systemic and mucosal sites(17, 18). Comparisons of OPV and IPV-induced immune responses are limited due to the widespread global use of OPVs which made the availability of completely OPV-naïve cohorts historically limited to just a few geographic regions. Herein, we leverage two distinct polio vaccine trials to examine differences in T cell immune responses following either an IPV-based (n=29) or monovalent OPV-based (n=28) vaccine dose (boost) in subjects who were either OPV or IPV-primed in childhood. Using a flow cytometric activation induced cell marker (AIM)-based approach we assessed the frequency and magnitude of CD4+ and CD8+ T cell responses to structural and non-structural viral proteins. T cell responses were then correlated with fecal viral shedding in OPV-boosted cohorts to begin to explore the role of cellular immunity in effective mucosal immunity to poliovirus.

## Results

### Characteristics of vaccine cohorts (**Fig. 1**)

We assessed Ag-specific T cell responses in a total of 57 healthy adult volunteers enrolled in two polio vaccine trials at the University of Vermont (UVM) Vaccine Testing Center. All volunteers had received complete polio vaccine series in childhood with either OPV or IPV-only based regimens. Characteristics of these cohorts are summarized in **Fig. 1** and **Table S1**.

**Figure 1:**
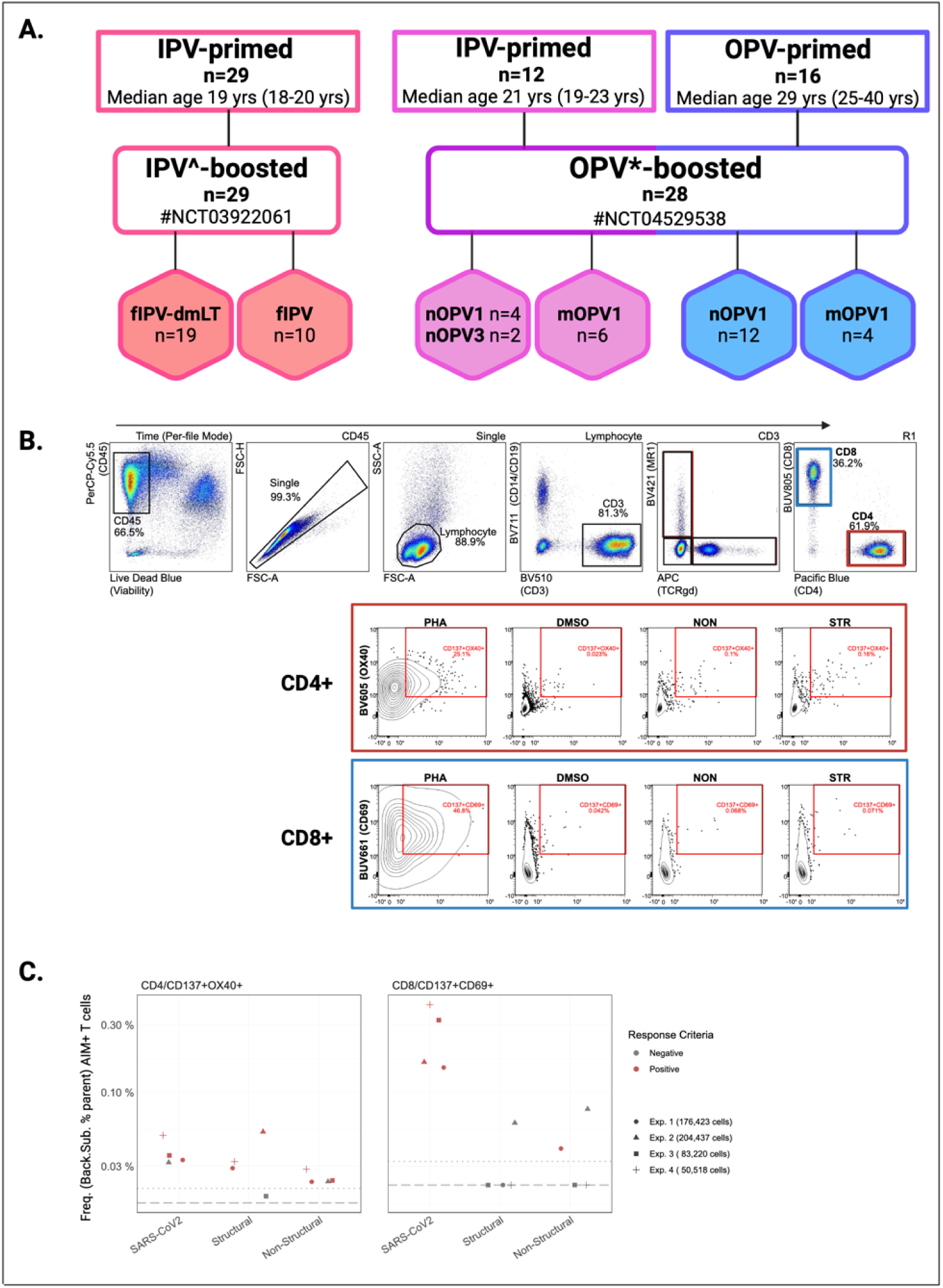
Polio vaccine study cohorts and detection of antigen-specific T cells by AIM+ assay. **A.** Volunteers (n) were enrolled in one of two polio vaccine studies conducted at the University of Vermont Vaccine Testing Center. In the IPV^-boosted study, IPV-primed healthy adults were randomized to receive an intradermal fractional dose of tIPV with or without dmLT. In the OPV*-boosted study, IPV-primed and OPV- primed cohorts were randomized to receive a novel or monovalent Sabin oral polio vaccine (OPV) strain for either serotype 1 or 3. **B.** Representative graphs depicting the flow cytometry gating strategy for defining poliovirus-specific CD4+ and CD8+ T cells by CD137+OX40+ and CD137+CD69+ expression, respectively. **C**. PBMCs from a single donor were included in each experimental run as an experimental control. Donor controls were stimulated with PHA, SARS-CoV2, and poliovirus (structural, and non-structural) peptide pools. Frequency (%) of AIM+ (CD137+OX40+) CD4+ T cells (left) and AIM+ (CD137+CD69+) CD8+ T cells (right) are shown. Each shape represents a separate experimental run (1–4) with the mean number of cells captured each experimental condition listed. Shapes are colored by response criteria (Background Subtraction > LOS; SI > 2). PHA, phytohemagglutinin; DMSO, dimethyl sulfoxide; MR1, MHC-I related molecule 1 (MAIT cell marker), TCRgd, T cell receptor gamma delta; NON, nonstructural peptide pool; STR, structural peptide pool; LOS, Limit of Sensitivity; SI, Stimulation Index ^IPV formulations include trivalent inactivated polio vaccine (tIPV) with or without dmLT (double mutant Labile Toxin); *OPV formulations include novel OPV serotype 1 (nOPV1); monovalent OPV serotype 1 (mOPV1); novel OPV serotype 3 (nOPV3), monovalent OPV serotype 3 (mOPV3)

Twenty-nine (N=29) of the volunteers included in this immunologic sub-study were enrolled in a single site Phase I polio vaccine trial (NCT03922061) in which healthy adults were randomized 2:1 to receive Sanofi’s licensed IPOL trivalent inactivated polio vaccine (tIPV; NDC 49281860-78) delivered intradermally at the dose-sparing fractional volume of 1/5 the full dose (0.1 mL) with or without 0.47 ug of a novel mucosal adjuvant, double mutant Labile Toxin (dmLT (LT(R192G/L211A)), a protein toxoid derived from wild-type enterotoxigenic *Escherichia coli* (ETC) labile toxin (LT). All volunteers had received only IPV-based polio vaccines in childhood. The median age across the study was 19 years (range 18 to 20); 72.4% were female and 96.6% of participants were white. Serum and peripheral blood mononuclear cell (PBMC) samples were collected at baseline and days 7, 10, and 28 following vaccine boosting. Serotype specific serum neutralizing antibody (SNA) titers were assessed at each time point, results of which are summarized in **Table S2;** complete trial results are published separately(19). Data and PBMCs collected at baseline, and days 7 and 10 following vaccination from all participants are included in the current analyses. Volunteers from this study are herein considered in the ‘IPV-primed/IPV-boosted’ group. (**Fig. 1**).

The remaining twenty-eight (N=28) volunteers included in this immunologic sub-study were enrolled in a multi-site Phase I polio vaccine trial (NCT04529538). This parent study randomized a total of 205 healthy adult volunteers to receive either a Sabin or novel monovalent oral live attenuated vaccine (OPV) strain. Vaccine groups included both exclusively IPV-primed and OPV-containing cohorts. The OPV vaccines administered in the parent trial included monovalent Sabin strain (mOPV1; 10^6.0^CCID50/0.5mL), monovalent novel (nOPV1; 10^6.5^CCID50/0.5mL), monovalent Sabin strain (mOPV3; 10^5.8^CCID50/0.5mL), monovalent novel (nOPV3; 10^6.5^CCID50/0.5mL) (Bio Farma (Indonesia)) and OPV-containing cohorts received a second dose of the experimental vaccine to which they were initially randomized on day 28. Notably, no volunteers randomized to receive monovalent Sabin strain (mOPV3) are included in this immunologic sub-study and only data and samples following the first vaccine dose up until Day 28 are included. Stool samples were collected from all participants on days 0, 7, 14, 21, 28 post initial dosing, and additionally on days 2, 4, and 9 for the IPV-primed cohort only. Stool sampling continued weekly until shedding cessation was confirmed by two consecutively negative stool samples. Serum and PBMC samples were collected at baseline and days 7 and 28 following vaccine boosting. Serotype specific serum neutralizing antibody (SNA) titers were assessed at baseline and day 28; complete results from this study will be reported separately (manuscript in submission). A subset of twenty-eight (n=28) volunteers enrolled at UVM and who consented to provide additional blood samples are included in the current analyses; SNA and fecal viral shedding data for these volunteers were provided by the study sponsor (**Table S2, S7**). Twelve of the 28 volunteers included herein received exclusively IPV in childhood and are herein considered in the ‘IPV-primed/OPV-boosted’ group (median age 21 years (range 19 to 23)) and the remaining 16 volunteers were OPV-only vaccinated in childhood and are considered in the ‘OPV-primed/OPV-boosted group’ (median age 29 years (range 25 to 40) (**Fig. 1**). Across both priming cohorts, 53.6% of the 28 participants were female and 96.4% identified as white (**Table S1**).

### Vaccine-induced adaptive immunity to polio includes both CD4+ and CD8+ T cell responses (Fig. 2A-C)

To investigate the impact of polio vaccines on adaptive cellular immune responses, we used PBMCs collected before and after vaccination to assess Ag-specific T cell responses using an activation-induced marker (AIM) assay in which cells were stimulated with poliovirus peptide megapools (MPs) of overlapping 15-mers by 10 amino acids. MPs were inclusive of all 3 poliovirus serotypes. Structural (n = 293) and non-structural (n = 368) peptides were separately synthesized and pooled. The frequency (% parent) of antigen(Ag)-specific CD4+ and CD8+ T cells were calculated per sample using a background subtraction method and only samples that met response criteria (background subtracted value (%) > limit of detection (LOD) and > limit of sensitivity (LOS), and stimulation index (SI) >2) were considered positive(20).

**Figure 2:**
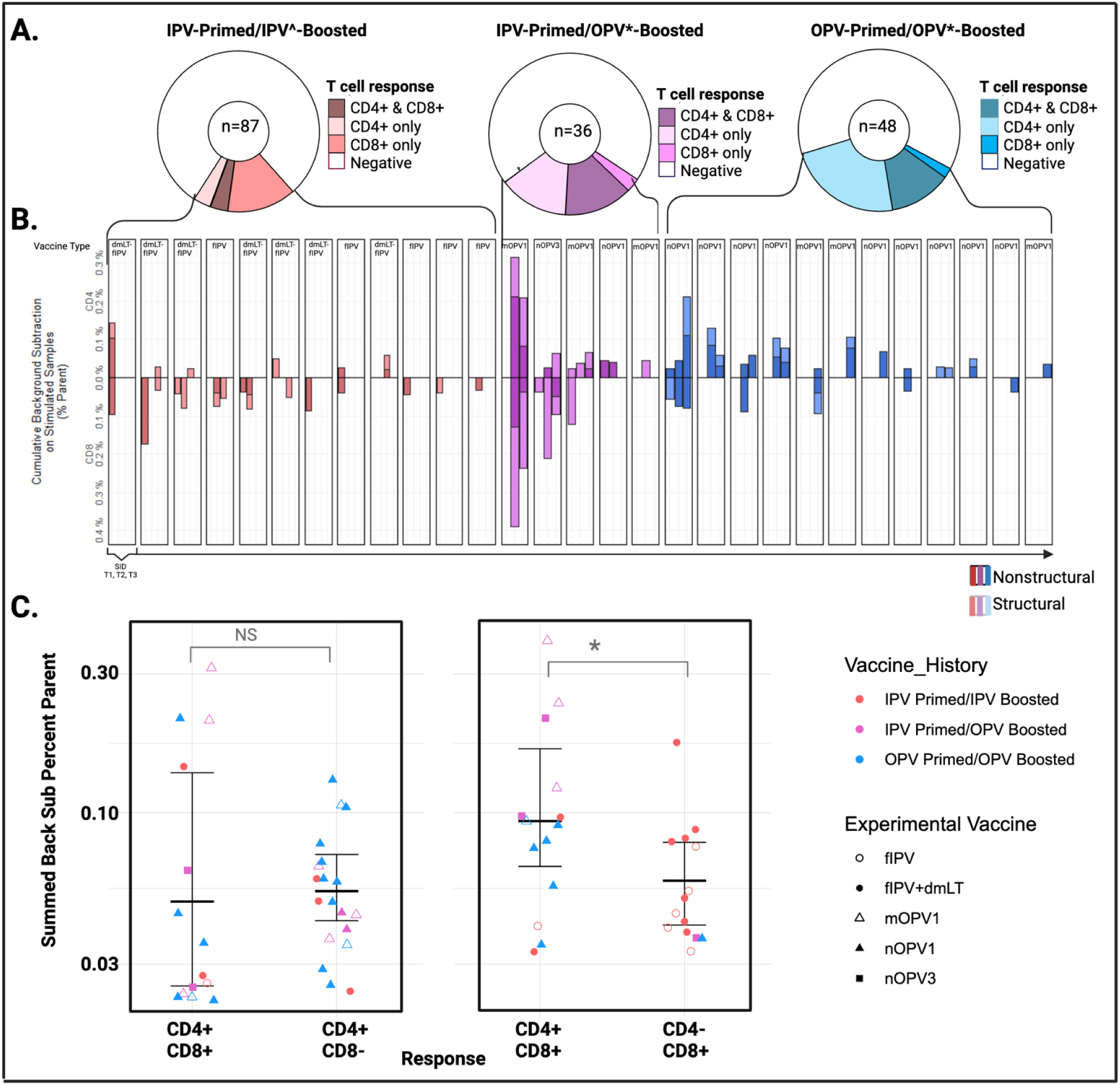
Overlap of CD4+ & CD8+ T cell responses per volunteer. **A.** The proportion of samples (n) with concurrent CD4+ and CD8+ T cell responses detected, those with only CD4+ or CD8+ T cell responses, and those that did not meet response criteria (negative). **B.** Total frequency of AIM+ (CD137+OX40+) CD4+ and (CD137+CD69+) CD8+ AIM+ T cells detected per sample. Total AIM+ frequencies are summed (structural (striped) and nonstructural (solid)) background subtracted values. Samples are arranged by subject and time point. **C.** Correlation of the magnitude (frequency (%)) of CD4+ and CD8+ T cell responses detected per sample. Values are summed and background subtracted. Points are shaped by epitope reactivity and colored by prime/boost group. IPV^ formulations include tIPV with or without dmLT; OPV* formulations include nOPV1, mOPV1, nOPV3, mOPV3; SID, subject ID; NON, nonstructural; STR, structural

As this was our first use of an AIM-based method to detect Ag-specific T cell responses to polio vaccines, we wanted to first investigate the prevalence, frequency, and overlap of polio Afumg-specific CD4+ and CD8+ T cells in our sample set. Across all cohorts, 38.6% (22/57) of volunteers had Ag-specific CD4+ and 33.3% (19/57) had Ag-specific CD8+ T cells detected in at least one sample, for a total of 33 samples meeting all response criteria for CD4+ and 28 samples meeting all response criteria for CD8+ T-cells (**Table S4)**. Fourteen of the positive samples had concurrent CD4+ and CD8+ Ag-specific T cells detected, 19 had CD4+ only and 14 had CD8+ only T cells detected. The distribution and overlap of CD4+ and CD8+ positive samples according to both study and subject are shown in **Fig. 2 A, B**. The mean frequency of Ag-specific T cells detected per sample was 0.07% of CD4+ T cells (range 0.023-0.315%) and 0.091% of CD8+ T cells (range 0.033-0.391%) (**Table S3**). Samples with concurrent CD4+ and CD8+ Ag-specific T cells (n=14) had higher frequencies of Ag-specific CD8+ T cells (mean frequency 0.12% of CD8+ T cells) compared to those (n=14) with only CD8+ T cells detected (mean frequency 0.063% of CD8+ T cells) (p = 0.04) (**Fig 2C**)

To assess the absolute magnitude of vaccine-induced T cell responses, we were able to calculate absolute cell counts using paired complete blood count (CBC) values for volunteers enrolled in the OPV-boosting vaccine study (NCT03922061). (**Table S6**) CBC testing was performed in the clinical laboratory at the University of Vermont Medical Center (UVMMC) using routine hematology analyzers. The mean absolute Ag-specific cell count calculated across all CD4+ reactive samples was 0.63 cells/mm^3^ (range 0.15 to 3.82 cells/mm^3^), with the highest mean counts detected in IPV-primed/OPV-boosted volunteers at Day 28 (1.23 cells/mm^3^; range 0.65-2.35 cells/mm^3^) **(Table S6)**. Compared to absolute Ag-specific CD4+ T cell counts, mean absolute Ag-specific CD8+ T cell counts were lower overall (0.36 cells/mm^3^ (range 0.08 to 1.48 cells/mm^3^)) and the highest mean levels were detected at Day 7, also in IPV-primed/OPV-boosted volunteers (0.91 cells/mm^3^ (range 0.62-1.21 cells/mm^3^) (**Table S6)**. To consider the impact of such small cell frequencies on future experimental designs, we calculated the expected number of Ag-specific T cells present in a standard 8mL blood draw tube and found that, on average, the total expected number of Ag-specific CD4+ and CD8+ T cells was 5,057 cells/8mL and 2,890 cells/8mL, respectively.

### Vaccination with oral live attenuated polio vaccines induce enhanced CD4+ T cell responses relative to inactivated formulations (Fig 3A-F)

CD4+ T cell responses (defined as post-vaccination detection of Ag-specific CD4+ T cells) were more frequently detected in OPV-primed/OPV-boosted (68.75%; 11/16) and IPV-primed/OPV-boosted (41.66% (5/12)) volunteers compared to those who were IPV-primed/IPV-boosted (10.35% (3/29); p < 0.0005) (**Fig 3C**). Ag-specific CD4+ T cells increased from baseline to Day 7 following OPV boosting in both priming cohorts (mean frequency of CD4+ T cells in IPV-primed: 0.00849% (Day 0) to 0.0164% (Day 7), p = 0.18; OPV-primed: 0.00688% (Day 0) to 0.0164% (Day 7), p = 0.01) (**Fig 3A**). Despite detection of some baseline positivity, which we believe may reflect higher cell numbers interrogated in samples from this cohort (**Fig. 1C**), a rise in Ag-specific CD4+ T cells was not detected in the IPV-primed/IPV boosted group until Day 10. Comparisons at these early time points (Day 7 following OPV boosting versus Day 10 following IPV boosting) revealed higher levels of Ag-specific CD4+ T cells in OPV-boosted groups, regardless of priming history (p=0.0192, IPV-primed/IPV-boosted vs. IPV-primed/OPV-boosted; p=0.0087, IPV-primed/IPV-boosted vs. OPV-primed/OPV-boosted)) (**Fig. 3A**). The highest mean frequencies of Ag-specific CD4+ T cells were detected 28 days post vaccination in the IPV-primed/OPV-boosted group (0.112% (range 0.063% to 0.208%) (**Table S3**); Day 28 samples were unavailable from the IPV-primed/IPV-boosted group for comparisons of later timepoints.

**Figure 3:**
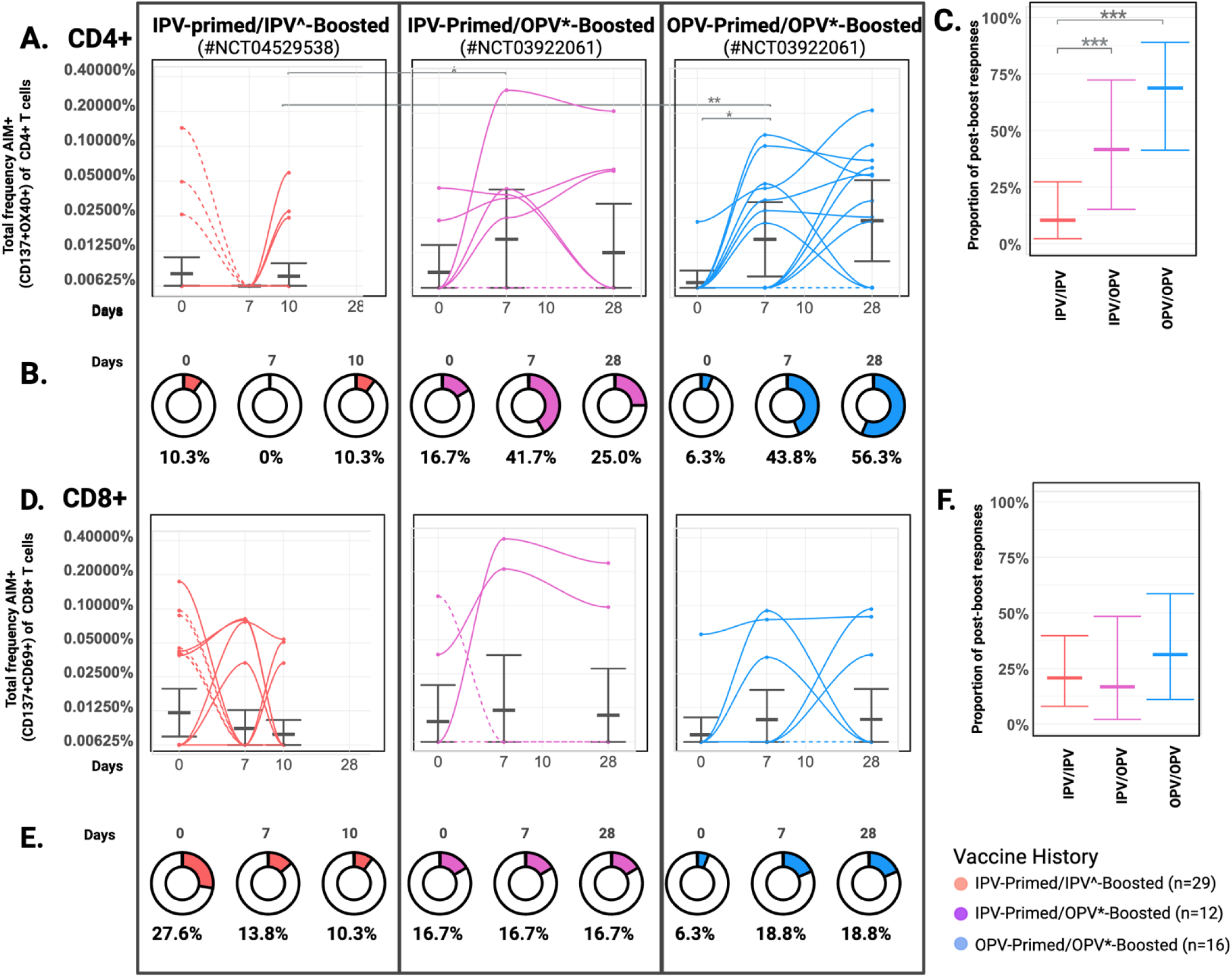
Oral live attenuated polio vaccines induce enhanced antigen-specific T cell responses relative to inactivated vaccine formulations. Poliovirus-specific T cells before and after boosting with an inactivated (IPV^) or oral live attenuated (OPV*) vaccine formulation. PBMC samples from baseline (Day 0) Days 7 and 10 in IPV^-boosted (n=29), and Days 7 and 28 in OPV*-boosted (n=28) adults were tested. Total poliovirus-specific T cell frequencies are provided as the summed percentage of AIM+ (CD137+OX40+) CD4+ or AIM+ (CD137+CD69+) CD8+ T cells detected after stimulation of PBMCs with structural and nonstructural peptide pools. Data are background subtracted against DMSO negative controls. **A., D**. Kinetics of CD4+ and CD8+ T cell responses per subject over time. Dashed lines indicate detection of Day 0 response only or no response. Geometric mean values are indicated with error bars indicating upper bound of the 95% CI. Statistical comparisons by Wilcoxian Rank Sum Test with p values <0.05 are indicated (*) using a false discovery rate (q value < 10%) to correct for multiple comparisons. **B., E.** Doughnuts represent the proportion of subjects with poliovirus-specific T cells detected at each timepoint. **C., F.** Proportion of volunteers with T cell responses detected following vaccination (not inclusive of baseline only detection) with error bars indicating upper bound of the 95% CI. Data is colored according to prime/boost vaccine group. IPV^ formulations include tIPV with or without dmLT; OPV* formulations include nOPV1, mOPV1, nOPV3; CI, confidence interval using Klopfer Pierson method; LOD, limit of detection.

CD8+ T cell responses (defined detection of Ag-specific CD8+ T cells in post-vaccination samples) were observed in a similar proportion of volunteers across groups (31% (5/16) of OPV-primed/OPV-boosted volunteers, (17%; 2/12) of IPV-primed/OPV-boosted, and 21% (6/29) of IPV-primed/IPV-boosted volunteers (**Fig 3F**), and mean frequencies were not statistically different between groups or timepoints (**Fig 3D**). The highest CD8+ T cell frequencies were detected IPV-primed/OPV-boosted volunteers on Day 7 (mean 0.301% of CD8+ T cells, range 0.211 to 0.391%) (**Table S3**); levels remained elevated in these individuals at Day 28 (**Fig 3D**). Again, Day 28 samples were unavailable from IPV-primed/IPV-boosted volunteers for comparisons of later timepoints.

### CD4+ T cell responses are associated with boosting of heterotypic serum neutralizing antibodies (Fig. 4)

To investigate the correlation between cellular and humoral adaptive immune responses, we assessed the relationship between Day 28 serotype-specific SNA titers and CD4+ T cell responses (**Fig. 4A**). OPV-primed volunteers boosted with oral monovalent vaccine formulations (#NCT03922061; PV1 or PV3 OPV strain) with CD4+ T cells showed increased Day 28 heterotypic antibody levels (mean titer 8.36 log2) compared to those without CD4+ T cells detected (mean titer 7.69 log2, p = 0.01). A similar difference was not observed in the IPV-primed/OPV-boosted group (**Fig 4B**). Volunteers in the IPV-primed/IPV-boosted group (#NCT04529538) received trivalent IPV (tIPV) formulations and exhibited universally strong SNA responses to all three polio serotypes at Day 28.

**Figure 4:**
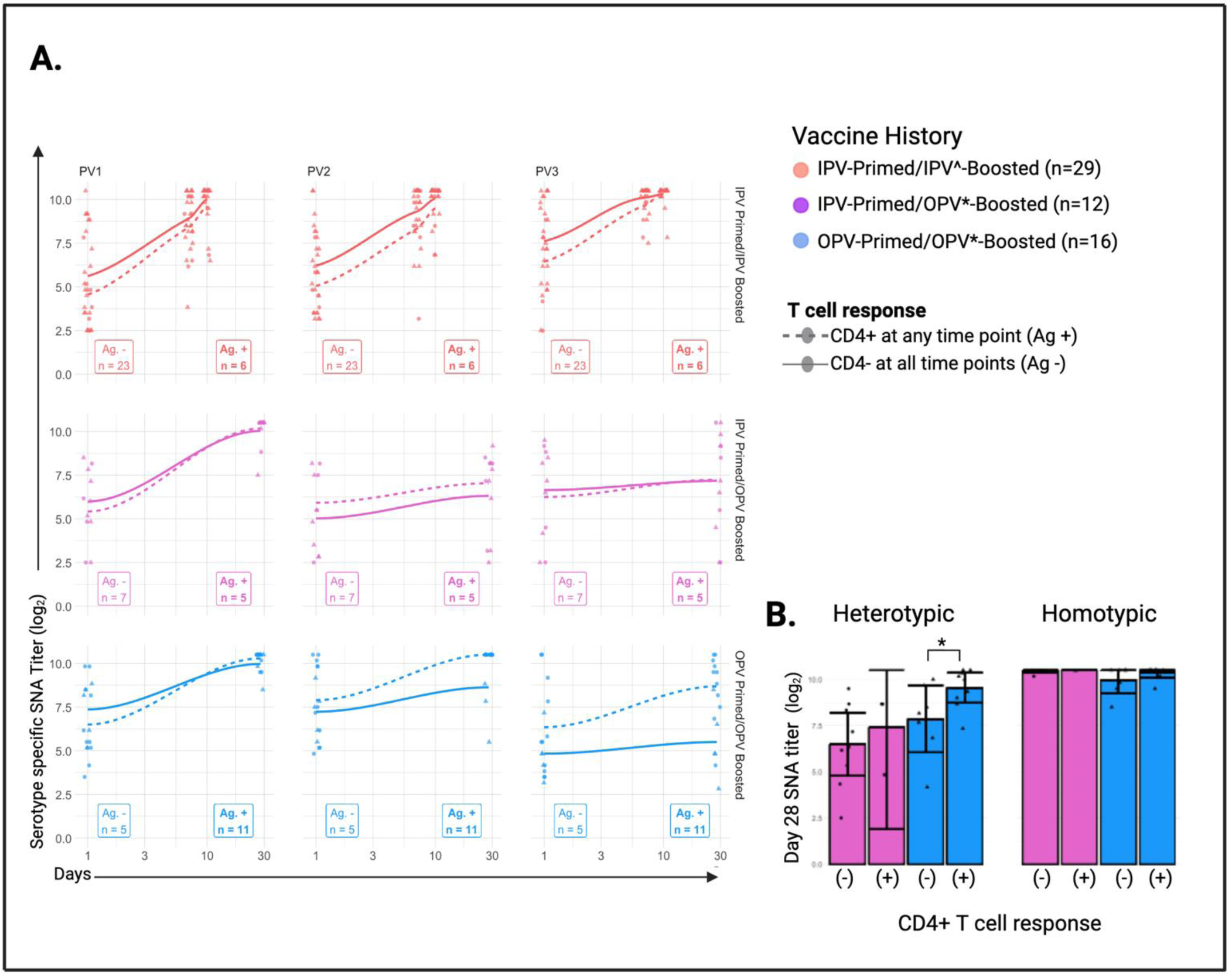
CD4+ T cell Responses and Day 28 Serum Neutralizing Antibody Titers. **A.** Mean SNA titers at baseline and day 28 following vaccine boosting for subjects with Ag-specific CD4+ T cells detected at any time point (dashed line) and those without Ag-specific CD4+ T cells (solid line). The number of subjects (n) included for each group is provided. **B.** Homotypic SNA titers to the matching viral serotype as the vaccine received. Heterotypic SNA titers are an average of day 28 titers to PV2 and remaining serotype (PV1 or 3) not received. Statistical comparisons made by Welch’s two sample t tests using a false discovery rate (q value < 5.5%) with p values < 0.05 indicated (*). IPV^ formulations include tIPV with or without dmLT; OPV* formulations include nOPV1, mOPV1, nOPV3, mOPV3; SNA, serum neutralizing antibody titer; PV1, poliovirus serotype 1; PV2, poliovirus serotype 2; PV3, poliovirus serotype 3

### Priming with oral live attenuated polio vaccines is associated with T cell recognition of non-structural viral proteins (Fig 5A-B)

To begin to explore the antigen specificity of T cell responses to polio vaccines and the impact of vaccine-priming on epitope responses we compared the relative proportion of Ag-specific T cells reactive to structural compared to non-structural peptides in each sample and across prime/boost groups.

**Figure 5:**
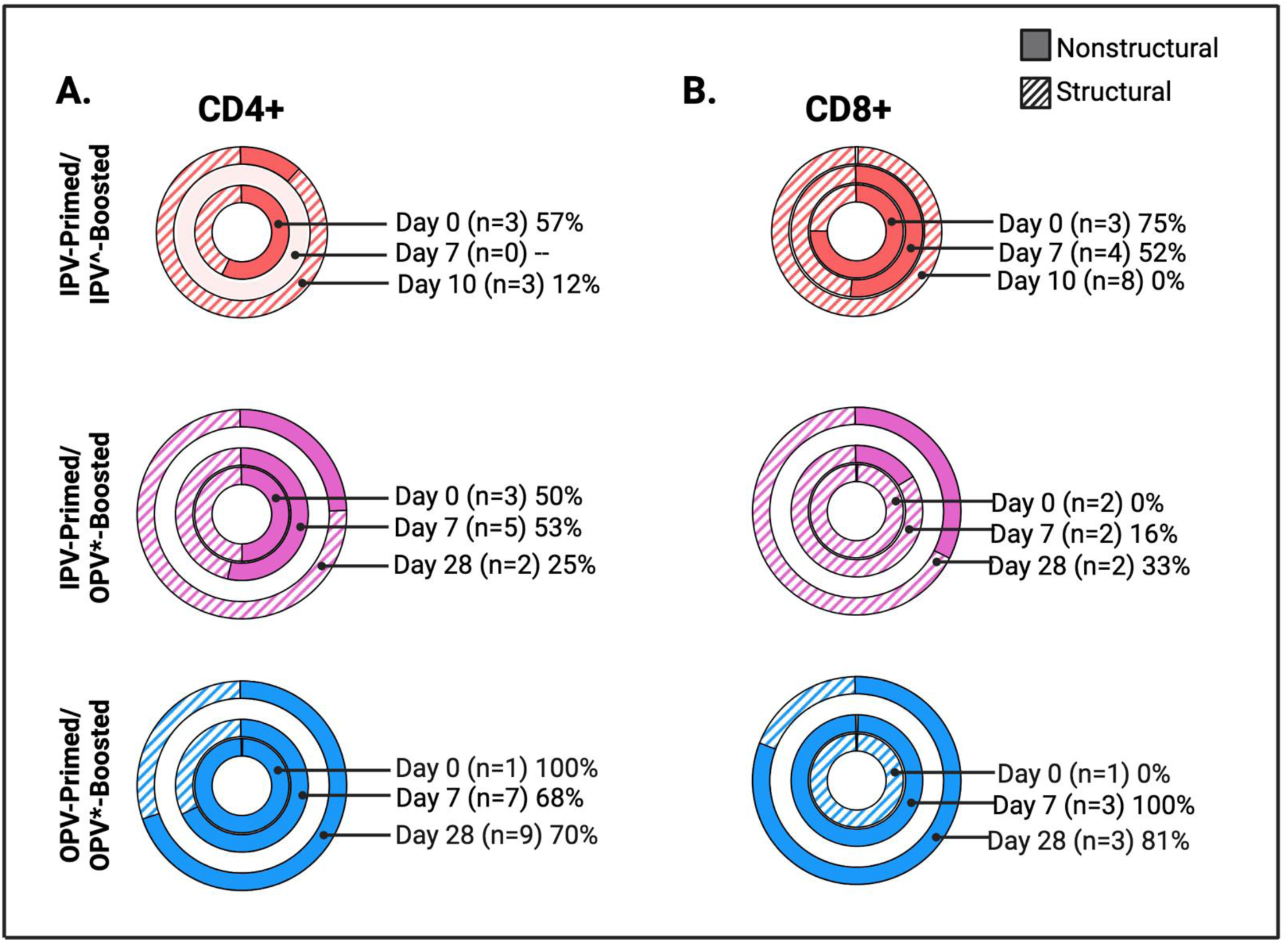
Priming with Oral Live Attenuated Polio Vaccines is associated with T cell recognition of non-structural viral proteins. The proportion of AIM+ T cells detected following stimulation with nonstructural (solid) and structural (striped) peptide pools relative to the total frequency of poliovirus-specific CD4+ **(A)** or CD8+ **(B)** T cells was calculated for each sample that met response criteria. The mean relative contribution (%) of non-structural epitope reactivity across samples (n) that met response criteria is listed for each time point.

Immunodominance was determined at the group level by calculating the overall proportion of T cell responses reactive to non-structural antigens within each prime/boost group at each timepoint (**Fig 5**). Samples responding to structural antigens alone were denoted as 0%. Strikingly, CD4+ and CD8+ T cell responses in OPV-primed/OPV-boosted volunteers were predominately to non-structural peptides (**Fig.5A,B**). This was most pronounced for CD8+ T cell responses in which 100% of the Ag-specific CD8+ T cells identified in Day 7 (n=3) and 81% in Day 28 (n=3) samples were to non-structural peptide pools. The proportion of the total CD4+ T cell response directed toward non-structural peptides was 68% (n=7) and 70% (n=9) in OPV- primed/OPV-boosted volunteers at days 7 and 28, respectively.

In contrast, an expansion of CD4+ and CD8+ T cell populations with specificity for structural peptides was more frequently observed in IPV-primed/OPV-boosted volunteers, with only 25% of their total CD4+ and 33% of their total CD8+ T cell responses directed to non-structural peptides at days 7 and 28, respectively (**Fig.5A, B**). The proportion of CD4+ and CD8+ T cell responses recognizing non-structural viral peptides was lowest in the IPV-primed/IPV-boosted group (12% of CD4+ (n=3) and 0% of CD8+ (n=8) T cell responses at Day 28). In comparing these groups, the difference in the proportion of Day 28 CD4+ T cell response directed toward non-structural peptides was statistically higher in the OPV-primed/OPV-boosted (70% (n= 9) compared that of the IPV-primed/OPV-boosted group (25% (n=2) (**Fig. 5A**). Although not as frequent, the proportion of Day 28 CD8+ T cell responses directed toward non-structural epitopes was also higher in the OPV-primed/OPV-boosted group (81% (n= 3) compared to that of the IPV-primed/OPV-boosted group (33% (n=2)) (**Fig. 5B**) suggesting that differences in vaccine priming may impact the immunodominance of subsequently expanded memory T cell populations and that enhanced recognition of non-structural proteins is associated with initial OPV priming.

At the individual sample level, 9 of the 33 samples with Ag-specific CD4+ T cells detected were reactive to structural antigens alone, 12 samples were reactive to non-structural antigens alone, and another 12 samples reacted to both structural and non-structural antigens (**Fig. S1A, Table S4**). Of samples with mixed structural/non-structural epitope responses, reactivity to non-structural proteins made up an average of 54.13% of the total CD4+ T cell response per sample (range 35.55% to 70.83%) (**Fig. S1C, Table S5**). Samples with CD4+ T cells reactive to both structural and non-structural peptides had statistically higher total Ag-specific CD4+ T cell frequencies detected (0.1273%) compared to those reactive to either non-structural (0.0372%; p = 0.003) or structural peptides alone (0.0360%, p = 0.002). This suggests to us that increased detection of these rare cell populations likely enhances observations of mixed epitope responses, as opposed to the existence of truly biologically exclusive epitope repertoires.

Of the 28 total samples with Ag-specific CD8+ T cells, 10 were reactive to structural antigens alone, 12 to non-structural antigens alone, and 6 reacted to both structural and non-structural antigens (**Fig. S1B, Table S4**). Similar to CD4+ T cell observations, samples with CD8+ T cell responses to both structural and non-structural proteins had higher total Ag-specific CD8+ T cell frequencies (0.1630%) compared to those reacting to only structural (0.0725%) or non-structural (0.0694%) peptide pools, although this difference was not statistically significant (mixed responses vs. structural only (p=0.14); vs. non-structural only (p=0.13)). In CD8+ reactive samples with mixed epitope responses, reactivity to non-structural epitopes made up on average 41.05% of the total CD8+ T cell response (range 32.86% to 53.05%) (**Fig. S1D, Table S5**).

### Duration and magnitude of OPV vaccine shedding are higher in IPV-primed compared to OPV-primed volunteers (Fig 6A)

To investigate the role of adaptive T cell responses in control of viral shedding, we assessed differences in fecal shedding following OPV boosting in volunteers enrolled in study NCT03922061 (“OPV-boosted”) as a function of both vaccine priming and T cell response. Rates, duration, and magnitudes of fecal vaccine shedding were measured in longitudinally collected stool samples by serotype-specific rt-PCR and positive samples quantitatively titered by traditional cell culture methods. For each volunteer, shedding cessation was confirmed by 2 consecutive stool samples testing negative for the presence of viral RNA by rt-PCR. The proportion of vaccinees with viral RNA detected and the arithmetic mean titer were calculated separately for OPV- and IPV-primed groups at each time point (**Table S7**). Additionally for each volunteer, time to shedding cessation and a shedding index endpoint (SIE) were calculated (**Table S8**). Time to shedding cessation was recorded as the number of days post OPV dosing that corresponded with the collection date of the first negative stool sample. Individual level SIE values were calculated as the mean viral titer observed in samples collected from a given volunteer on Days 7, 14, 21, 28. Mean SIE values were compared between groups.

**Figure 6:**
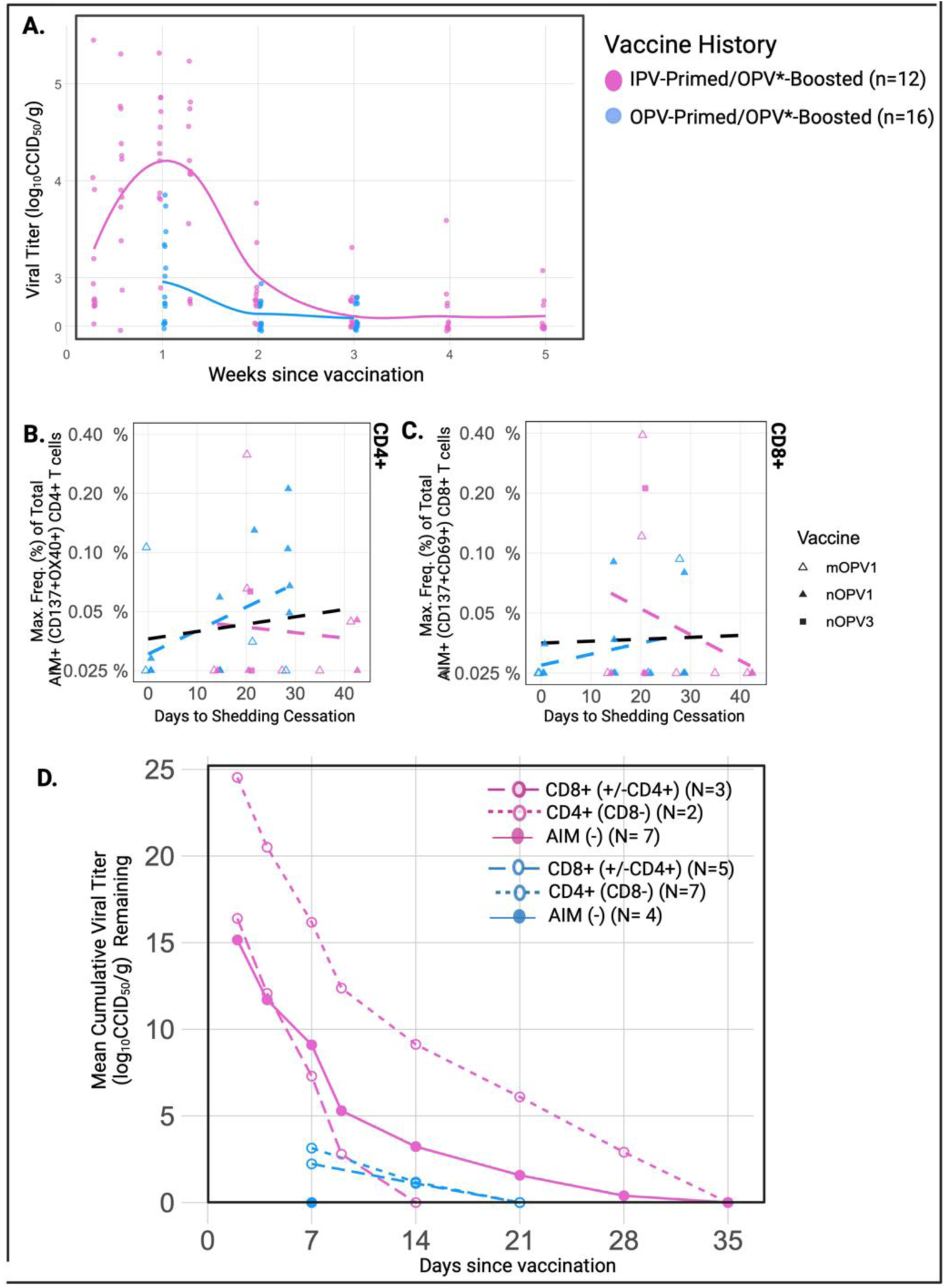
Primary CD8+ T cell responses are associated with control of viral shedding. Stool samples were longitudinally collected from subjects enrolled in the OPV*-boosting study (#NCT03922061) to measure rates and amounts of fecal shedding of OPV vaccines. Shedding cessation was confirmed in all subjects by two consecutive stool samples testing negative by rt-PCR. **A**. Viral titer levels of stool samples positive by rt-PCR. **B-C,** Correlation of time to shedding cessation and maximum frequency of total AIM+ (CD137+OX40+) CD4+ **B.** and AIM+ (CD137+CD69+) CD8+ **C.** T cells detected per subject. Regression lines provided for all samples (black) and for each prime/boost group. **D.** The mean cumulative viral titer remaining over time for volunteers with and without CD8+ T cell responses by priming cohort. OPV* formulations include nOPV1, mOPV1, nOPV3, mOPV3CCID50, 50% cell culture infective dose; SIE, Mean shedding index endpoint; Max., maximum; Freq., frequency

At all shared timepoints, higher rates of fecal shedding as well as higher mean viral titers were detected in the IPV-primed compared to OPV-primed group. At Day 7, for example, viral shedding was detected in all (100%) IPV-primed vaccinees (mean titer 4.24 log10CCID50), compared to 68.8% of OPV-primed individuals (mean titer 3.15 log10CCID50) (**Fig. 6A**, **Table S7**). The mean time to shedding cessation was also significantly longer in IPV-primed individuals (26.833 versus 14.875 days (p< 0.0001)) (**Table S8**). Mean SIE, which accounts for both an individual’s magnitude and duration of viral shedding, was also significantly higher in the IPV-primed compared to OPV-primed group (2.229 versus 1.063, p<0.0005) (**Table S8**). Notably, of the 31% (5/16) of volunteers with no viral shedding detected at Day 7, all were OPV-primed (**Fig. 6A, Table S7**). A maximum time to shedding cessation of 42 days was observed in three (n=3) IPV-primed volunteers) (**Fig. 6A, Table S8**).

### Primary CD8+ T cell responses are associated with control of viral shedding (**Fig 6B-D**)

Fecal OPV shedding provides a surrogate marker of effective Ag-specific mucosal immunity to poliovirus(21). To investigate the role of T cells in mucosal control of viral shedding, we correlated the maximum total frequency (% parent gate) of CD4+ and CD8+ T cell responses observed for each volunteer with their time to shedding cessation and SIE values (**Fig. 6B-C**). Due to the known impact of vaccine priming on OPV shedding dynamics, statistical comparisons were limited to within each vaccine priming cohort. We also limited formal statistical comparisons to those with or without any T cell response (inclusive of any combination of CD4+ and CD8+ T cell response) and did not further subdivide by antigen specificity due to our relatively small sample size (**Table S9**).

The relationship of peripheral T cell responses with vaccine shedding dynamics appear to be different in OPV-compared to IPV-primed cohorts. When describing these relationships, it is important to remember that OPV replication is expected to elicit a tissue-based memory response in OPV-primed volunteers while OPV replication in this cohort of IPV-primed individuals represents initial antigen exposure at mucosal sites. In total, CD4+ and/or CD8+ T cell responses were detected in 68.75% (11/16) OPV-primed volunteers. Either absent or low magnitude responses were detected in non-shedders and magnitudes generally increased with increasing time to shedding cessation (**Fig. 6B-C**) and SIE values (**Fig. S2**). Statistical comparisons revealed higher mean SIE values (1.30 versus 0.35, (p = 0.0156)); and longer shedding durations (17.50 versus 7 days, (p < 0.0001)) in OPV-primed volunteers with CD4+ and/or CD8+ T cell responses compared to those without T cell responses detected (**Table S9**). The proportion of total CD4+ T cell reactivity to non-structural proteins was higher in OPV-primed volunteers with shedding (72.2%-100% (n=13 samples) versus 43% (n=4 samples) in non-shedders) while the proportion of total CD8+ T cell reactivity to non-structural proteins was highest in non-shedders (100% (n=3 samples) than shedders (60.5% (n=4 samples)). (**Fig S3**) Taken together, this suggests a positive association exists between the amount and duration of viral replication in the gut of OPV-primed individuals and our ability to detect cellular immune responses in the peripheral blood.

Ag-specific T cells (CD4+ and/or CD8+) were identified in 42% (5/12) of IPV-primed volunteers and while the largest T cell responses were observed in IPV-primed individuals with the most robust viral shedding, responses were notably absent or diminished in those with the largest shedding durations (**Fig. 6B,C**) and SIE values (**Fig. S2**, **Fig. S3D,G**). Five IPV-primed volunteers continued to shed virus ≥ 21 days and despite robust boosting of Day 28 homotypic SNA titers (mean PV1 titer 10.43), T cell responses were generally absent in these individuals. None of the five IPV-primed volunteers with prolonged shedding had detectable CD8+ T cell responses at any timepoint and despite low levels of CD4+ T cell responses in two of the five at Day 7, none were detected at Day 28 (**Fig. 6B,C; Fig. S3**). Boosting of heterotypic SNA titers in this group were also minimal (Day 28 mean 5.4 (PV2); 5.5 (PV3)), suggesting lack of coordinated primary adaptive immune responses in IPV-primed individuals with prolonged fecal shedding.

The proportion of the overall T cell response directed towards nonstructural proteins was also lower in IPV-primed compared to OPV-primed shedders (34.4% (n=7 samples) of the CD4+ T cell response and only 16.3% (n=6 samples) of the CD8+ T cell response). (**Figure S3**) It is unknown if increased recognition of non-structural proteins is related to the relative impact of initial OPV versus IPV priming on the character of subsequently expanded memory T cell populations or if this is instead reflective of differences in the timing and character of a primary (in IPV primed) versus memory T cell response (in OPV primed) groups.

To further address our overarching hypothesis that OPV-induced activation of cytotoxic CD8+ T cells are important in controlling viral replication and shedding at mucosal surfaces, we calculated a shedding rate cessation curve in which cumulative mean viral titer amounts were calculated over time for volunteer groups with Ag-specific CD8+ T cells (+/- CD4+ T cells) and without Ag-specific CD8+ T cells (separate curves calculated for those with Ag-specific CD4+ T cells only and those without any detectable Ag-specific T cells) (**Fig 6D**). In total, Ag-specific CD8+ T cells were observed in 31% (5/16) of OPV-primed and 25% (3/12) of IPV-primed volunteers. Intriguingly, detection of Ag-specific CD8+ T cells in IPV-primed volunteers was associated with notably shorter durations of vaccine shedding (up to 14 days, compared to 35 days in those without Ag-specific CD8+ T cells). Alternatively, the shortest shedding durations observed in OPV-primed groups (<7 days) occurred in those without any Ag-specific T cells detected (n=4) and detection of CD8+ T cell responses was not associated with reduced shedding durations. (**Fig 6D**)

## Discussion

Here we have shown that live attenuated OPVs induce more frequent T cell responses compared to inactivated (IPV) formulations and that differences in vaccine priming in childhood strongly influence the breadth of T cell antigen specificity following subsequent polio vaccine exposure in adulthood. More specifically, OPV-priming is associated with expansion of T cell populations recognizing non-structural viral peptides while those observed in IPV-primed adults predominately react to structural viral peptides. We believe that differences in both the frequency and character of polio vaccine-induced T cell memory responses may help to explain why repeated vaccination with OPVs leads to further reductions in fecal shedding when compared to use of IPVs.

Data on the role of T-cell mediated cellular immunity to PV is sparse. While some reports have demonstrated CD4+ T cell responses in the peripheral blood of OPV-vaccinated individuals, data regarding the presence of CD8+ T cell responses is mixed and comparisons with OPV-naïve populations are limited due to the historically widespread use of OPVs (22–25). In addition, methodologic approaches employed in historical studies often lack detail regarding the type and character of T cell responses detected. Following removal of OPVs from infant vaccine schedules in the United States in the early 2000’s, the present study utilizes modern immunologic techniques to provide a unique comparison of T cell responses following boosting with a single dose of either an IPV (n=29) or OPV vaccine formulation (n=28) in both OPV-primed and OPV-naïve (IPV-primed) healthy adults.

Strikingly, we observed CD4+ T cell responses more frequently following OPV boosting compared to IPV boosting, with the highest proportion of responders detected in the OPV- primed/OPV-boosted group (68.75%; 11/16) (**Fig 3C**). Regardless of priming history, OPV boosting induced rapid expansion of Ag-specific CD4+ T cells by Day 7 and mean levels continued to rise until Day 28 sampling. The ability of live attenuated vaccines to induce superior T cell responses compared to inactivated vaccine formulations is observed for other viral pathogens, including influenza virus(26–28). In one study, generation of Ag-specific T cell responses correlated with protection against culture-confirmed clinical disease(29) and in another challenge model, recent exposure to live attenuated influenza vaccine (LAIV) reduced viral shedding compared to inactivated influenza vaccines (IIV) (30). Taken together, it appears that live attenuated vaccines induce robust pathogen-specific T cell responses capable of controlling viral replication, disease, and transmission. We speculate that that OPV-mediated induction of Ag-specific T cell responses may similarly help to control poliovirus shedding at sites of infection, such as the gut and oral pharynx.

Another major finding of this study is the difference in immunodominance observed in T cell responses following boosting with IPV compared to OPV vaccines and, perhaps more surprising the apparent retention of a predominately structural T cell response in IPV-primed volunteers despite recent boosting with an OPV vaccine. We believe this may reflect differences in immune priming and memory generation occurring during initial antigen exposure with inactivated IPVs compared to live attenuated OPVs. Replication of OPVs within mucosal tissues more closely mimics natural infection by providing antigenic exposure to the full suite of viral proteins expressed during viral replication, including to both structural and non-structural epitopes. Subsequent adaptive immune responses therefore include cross-reactive memory T cell populations with recognition of highly conserved non-structural viral proteins able to quickly recognize and kill virally infected cells upon reinfection. Enhanced T cell recognition of non-structural viral epitopes may therefore play an important role in OPV-induced mucosal immunity, as well as our observation of enhanced heterotypic neutralizing antibody responses in OPV- primed cohorts. While IPV-primed volunteers showed limited boosting of heterotypic neutralizing antibodies regardless of CD4+ T cell responses, detection of Ag-specific CD4+ T cells in OPV- primed volunteers was associated with enhanced boosting of heterotypic neutralizing antibodies (SNA levels to viral serotypes other than that with which they were boosted (Day 28 mean titer: 8.36 versus 7.69 log2, p = 0.01)). Interestingly, enhanced detection of heterotypic neutralizing antibody responses is described following natural infection with polioviruses or vaccination with OPVs but not following vaccination with IPVs or virus-like particles(31–33). This suggests that OPV-induced memory CD4+ T cell populations confer some amount of heterotypic immunity, potentially through T cell-dependent activation of cross-protective memory B cell populations capable of producing more broadly neutralizing antibody responses. The durability and clinical relevance of these heterotypic antibody responses, however, is unclear (34).

To investigate the potential relevance of T cell-mediated immunity in control of poliovirus shedding, we then sought to correlate T cell responses with fecal vaccine shedding dynamics following OPV-boosting. In keeping with prior literature, both shedding rates and amounts were markedly reduced in OPV-primed compared to IPV-primed volunteers, with all non-shedders being OPV-primed and the longest shedding durations (≥ 28 days) observed in IPV-primed volunteers. (**Fig. S3**) When examining the relationship between T cell responses and fecal shedding, notable differences were again observed between priming groups. While positive relationships were generally observed between the magnitude of T cell responses and vaccine shedding amounts (SIE) and durations, a notable lack of Ag-specific T cell responses were observed in IPV-primed volunteers with robust vaccine shedding of extended shedding durations (≥ 28 days). This observation was further highlighted when comparing shedding cessation curves of IPV-primed volunteers with and without Ag-specific CD8+T cells. All IPV-primed volunteers with detectable Ag-specific CD8+ T cells had stopped shedding by Day 14, compared to Day 35 in other groups. The same pattern was not observed in OPV-primed groups where reductions in shedding were instead linked to a lack of Ag-specific T cell responses. Taken together we believe that these differences likely reflect distinctive underlying biology occurring in OPV- compared to IPV-primed cohorts. We hypothesize that early control of viral shedding (<7 days) occurs via OPV-mediated induction of effective tissue-resident memory T cell populations that will not be detectable in the blood unless enough viral replication occurs that T cell populations are sufficiently boosted (and thus detectable). In IPV-primed cohorts without these tissue-resident memory populations however, viral replication will be allowed to occur with subsequent stimulation and trafficking of memory T cell populations to the gastrointestinal tract. Detection of Ag-specific CD8+ T cell populations in the peripheral blood of this group is therefore associated with control of viral replication, while a lack of Ag-specific T cells in the periphery of IPV-primed individuals signals a failure to mount an effective cytotoxic T cell response.

Virus-specific CD8+ T cells are shown to control replication and shedding of other mucosal viruses, including influenza, VZV, HIV, and SAR-CoV2 (35–38) and recent studies examining hetero-variant protection following natural infection with respiratory viruses have demonstrated the importance of cross-reactive CD8+ T cell populations, which can migrate between the blood and tissues (39–45). In a recent SARS-CoV2 challenge study for example, viral control was significantly correlated with induction of Ag-specific CD8+ T cell responses in cynomolgus macaques, even in the absence of neutralizing antibodies(35).

Fecal shedding of live attenuated OPVs sustains ongoing viral transmission and leads to the emergence of neurovirulent circulating vaccine-derived PV strains (cVDPVs)(6, 7). This study provides novel insights into the differential activation of CD4+ and CD8+ T cells following different prime/boost exposures to IPV and OPV vaccines and suggests a potential role for CD8+ T cells in control of poliovirus replication and shedding. While the live attenuated nature of OPVs likely support initial development of T cell responses, safer vaccine formulations without the risk of reversion could consider use of novel adjuvants or other innovative strategies aimed at reducing viral shedding and transmission through activation of CD8+ T cell populations.

## Limitations

This study has several limitations. Notably, the overall sample size is small and the generalizability of immunologic findings in adult populations to those most at-risk for poliomyelitis is limited. Use of fractional dose IPV delivered by intradermal injection with or without inclusion of a mucosal adjuvant (dmLT) in the IPV-boosted study also limits the overall generalizability of results given that intramuscular delivery of full dose non-adjuvanted IPV is common practice globally. Additionally, lack of day 28 samples in the IPV-primed/IPV-boosted cohort limits comparisons with OPV-boosted group and differences in age between IPV-primed and OPV- primed cohorts may confound immunologic outcomes. Additional studies in target populations with extended sampling durations would provide useful information on the overall kinetics and durability of vaccine-induced T cell responses to polio.

Similar to studies in other viral pathogens, we found Ag-specific T cells to represent very low frequencies of peripheral blood T cell populations, ranging from 0.02% to 0.39% of respective parent gates. Reliance on detection of these low frequency events using peripheral blood samples likely fails to capture the true breadth of T cell responses mediating mucosal immunity at barrier sites, including tissue resident memory T cells whose recirculation following repeated antigen stimulation can be difficult to capture with limited sampling intervals(46). Another potential limitation of using an AIM-based approach is identification of T cells cross-reactive with other closely related viruses, such as Coxsackieviruses or other enterovirus species. To increase the specificity of our findings, we employed conservative gating and response criteria however additional reproducibility studies (**Fig 1C**) and calculations of absolute cell counts using hematology analyzer data suggests that we are near the limit of detection of our assay, signaling an important challenge in balancing assay sensitivity and specificity and suggesting that our results may underrepresent the true prevalence of PV-induced T cell responses.

In conclusion, these data suggest that T cell responses, particularly cross-reactive CD8+ T cells with recognition of conserved non-structural viral proteins, may contribute to effective pathogen clearance and reductions in polio viral shedding. Novel vaccine strategies targeting these cell-mediated effector mechanisms may be of benefit in the final stages of the polio endgame. Future studies should plan to extend this work into pediatric populations, as they remain the prime target population for poliovirus vaccines and age-related differences in cellular immune function may be of critical importance.

## Data Availability

All data produced in the present study are available upon reasonable request to the authors.

## Author contributions

J.W.C., conceived of this work, oversaw project management, data generation, analysis, and significantly contributed to manuscript preparation.

B.D.K helped conceive of this work, provided scientific input, and significantly contributed to manuscript preparation.

A.G. helped to design and trouble-shoot Ag-specific T cell assay development, analyze Ag-specific T cell data, and provided significant conceptual and scientific input in manuscript preparation and writing.

A.S. provided support for the development of the AIM-based assay and generation of peptide megapools.

B.M. oversaw generation of serum neutralizing antibody and fecal viral shedding data, including molecular and cell culture-based assay approaches.

J.P.S. performed specimen processing, assay development and generated flow-cytometry-based immunology data, performed primary data analysis and contributed significantly to manuscript preparation.

P.H. performed additional data integration and analysis and generated figures.

R.G. aided in flow-cytometric assay development and generation of primary immunologic data sets.

M.S. generated figures for publication and significantly contributed to manuscript preparation.

## Acknowledgments

The authors are grateful to study participants for their contributions to the study. We thank members of the UVM Vaccine Testing Center and UVM Medical Center Clinical Research Center for their efforts in implementing the included studies. We thank PATH for the opportunity to use samples and additional metadata for the NCT04529538 study. We thank the laboratory staff that performed the serology testing (Basit Jafri, Kathryn Jones, William Hendley, Giovanna Sifontes, Sandra Valdez, and Yiting Zhang) and shedding testing (Talha Abid, Alex Alvarado, Elizabeth Coffee, Larin McDuffie, Esther Morantz, Lindsey Oteyza, Annelet Vincent, Ling Wei, and Amanda Williams) at the Polio and Picornavirus Branch and the Specimen Triage and Tracking Team (STTAT) at the Centers for Disease Control and Prevention (CDC). We are grateful to Drs. Dimitry Krementsov, Jon Boyson, and Sean Diehl for their helpful discussions and thoughtful input regarding flow cytometry panel design and data analysis throughout the development and implementation of this work. We thank The Harry Hood Bassett Flow Cytometry and Small Particles Detection (FCSPD) Core Facility at the UVM Larner College of Medicine. Figures made with Biorender.

This work was financial supported by the University of Vermont Department of Pathology and Laboratory Medicine and funded in part with Federal funds from the National Institute of Allergy and Infectious Diseases, National Institutes of Health, and Department of Health and Human Services under Award Numbers K23AI175660 (JWC), P30GM118228 (BK), and contract no.75N93024C00056 (A.G. and A.S). The content is solely the responsibility of the authors and the conclusions, findings, and opinions expressed by authors do not necessarily reflect the official position of the U.S. Department of Health and Human Services, the Public Health Service, the Centers for Disease Control and Prevention, or the authors’ affiliated institutions.

## Supplemental Methods

### Clinical trial details

Samples and data from human subjects included in the current study originated from participants enrolled in polio vaccine studies conducted at the University of Vermont Vaccine Testing Center (Burlington, VT (USA)). The first study (#NCT04529538) was completed in 2019 and complete trial results are previously reported(19). The study protocol was approved by the United State Food and Drug Administration’s (FDA) Investigational New Drug program (IND#18511) and by the Institutional Review Board (IRB) at the University of Vermont (UVM). This study was conducted in compliance with the ethical principles of the Declaration of Helsinki and all participants provided written, informed consent for use of their data and samples in future research.

The second multisite study (#NCT03922061) was completed in 2023, and complete results will be reported separately (manuscript currently under preparation). The study protocol for study #NCT03922061 was approved by the United State Food and Drug Administration’s (FDA) Investigational New Drug program (IND# 026305) Advarrra IRB and the University of Vermont (UVM) IRB. This study was conducted in compliance with the ethical principles of the Declaration of Helsinki and all participants provided written, informed consent. Participants enrolled at UVM were invited to participate in a separate substudy (STUDY0001320) to provide additional blood samples for the work presented here. Volunteers who agreed to substudy participation provided written, informed consent for use of their data and samples in future research.

Testing of serum and stool samples for both studies was reviewed by CDC, deemed research not involving human subjects, and was conducted consistent with applicable federal law and CDC policy (See e.g., 45 C.F.R. part 46; 21 C.F.R. part 56; 42 U.S.C. §241(d), 5 U.S.C. §552a, 44 U.S.C. §3501 et seq.)

### Serum neutralizing antibody titers

For both studies, (NCT04529538; NCT03922061), serum samples were collected for vaccine humoral immunogenicity assessments. For participants enrolled in study #NCT04529538, serum samples were collected on Day 0 prior to dosing, and on Days 7, 10, 14, and 28. For study #NCT03922061 serum samples were collected at the screening visit and on Day 28 for all cohorts. All samples were aliquoted and stored at ≤-20°C before being shipped to Polio and Picornavirus Branch at the United States Centers for Disease Control and Prevention (CDC) for assessment of type-specific polio neutralizing antibody titers(47). To enable comparison across studies, only results from baseline and day 28 samples are included in the current analysis.

### Poliovirus shedding

For study (NCT03922061), stool samples were collected on Days 7, 14, 21, 28 for all volunteers and additionally on Days 2, 4, and 9 for IPV-primed cohorts to detect and assess the quantity of virus shed and to confirm cessation of shedding. Samples were processed at the University of Vermont and stored at ≤-20°C before being transported to the Polio and Picornavirus Branch at the United States Centers for Disease Control and Prevention (CDC). The detection of type-specific poliovirus in stool was determined via multiplex real-time polymerase chain reaction (PCR). In samples positive by PCR, type-specific infectious virus was quantified as the 50% cell culture infectious dose (CCID50) per gram of stool.

The mean viral RNA titers and Shedding Index Endpoints (SIE) were calculated as the arithmetic mean across each group. The common time points (Days 7, 14, 21, and 28 post-vaccination) were used in the calculation of the mean SIE, which included zeroes for PCR negative observations. The arithmetic mean of time to shedding cessation (days) across each group were compared as opposed to a time-to-event analysis as the data were not censored; stool samples were continually collected until cessation of viral shedding was observed for each subject.

### Identification of Poliovirus-Specific T cell Reponses by Flow Cytometry Design of Polio Peptide Pools

To develop a pool of representative poliovirus peptides for research into poliovirus specific lymphocytes, polyprotein sequences available in the Virus Pathogen Database and Analysis Resource (ViPR) were analyzed for sequence redundancy. Nine strains available in ViPR were used in this analysis: two human poliovirus 1 strains Mahoney (GenBank Accessions: V01149, V01148), one human poliovirus 1 strain Mahoney_CDC (GenBank Accession: KU866422), three human poliovirus 1 strains Sabin 1 (GenBank Accessions: AY184219, V01150, GQ984141), one human poliovirus 2 strain MEF-1 (GenBank Accession: AY238473), one human poliovirus 2 strain Sabin 2 (GenBank Accession: AY184220), and one human poliovirus 3 strain Sabin 3 (GenBank Accession: AY184221). Per each sequence 15-mers overlapping by 10 amino acids were generated using the cluster tools used with 70% conservancy (PMID: 30014462) available in IEDB (PMID: 31114900). As a result, we identified a total of 661 peptides, 134 were uniquely represented in either of the strains while 527 were represented in one or more of the representative strains. The 661 peptides were synthetized by A&A (San Diego) as crude material. Peptide assignment to each antigen was performed as previously reported taking into account mapping positions across several Enteroviruses isolates including Polioviruses(48).

Peptides were resuspended in DMSO, pooled as structural (STR; n = 293) or nonstructural (NON; n = 368). To generate the two different megapools, the peptide pools followed a sequential lyophilization approach previously published(49). The resulting lyocake was resuspended in DMSO at a stock concentration of 1mg/mL generating the STR and NON megapools. As a control, a SARS-CoV-2 spike megapool (COV) was also utilized based on 15- mers overlapping by 10 amino acids spanning the spike SARS-CoV-2 ancestral strain (GenBank: MN_908947)(50). Megapools were generated by the Grifoni and Sette Labs at La Jolla Institute for Immunology and were maintained at -20 degrees Celsius with a maximum of one thaw prior to use in this assay.

### Isolation of Peripheral Blood Mononuclear Cells (PBMCs)

PBMCs were isolated by University of Vermont Vaccine Testing Center personnel according to standard procedures. Briefly, whole blood was collected from study participants in EDTA tubes and layered onto Histopaque-1077 (Sigma #H8889) for centrifugation. PBMCs were resuspended in cell freezing media (Sigma #C6164) and 6-10 million PBMCs were aliquoted per cryovial. Cryovials were frozen down in Mr. Frosty containers at -80 degrees Celsius and then transferred to liquid nitrogen for long-term storage.

### Thawing of Peripheral Blood Mononuclear Cells (PBMCs)

RPMI-1640 (Cytiva #SH30096.02) supplemented with heat-inactivated fetal bovine serum (“FBS”, Corning #35-011-CV, 10%), L-Glutamine (Gibco #A2916802, 1%), and Penicillin-Streptomycin (Gibco #15070063, 1%), was used as a base media throughout this protocol and is referred to as “RPMI-C.” Cryovials were removed from liquid nitrogen and placed in a water bath at 37 degrees Celsius to thaw for 60 seconds. One mL of sterile RPMI-C containing Benzonase (Sigma-Aldrich #E1014-25KU, working concentration 50 units/mL) was added to each cryovial and contents were briskly poured into conical tubes containing 3 mL sterile RPMI-C. Conical tubes were centrifuged at 1200 rpm for 10 minutes at 10 degrees Celsius and the supernatant was removed by gentle pouring. Cell pellets were resuspended in 1 mL RPMI-C and centrifuged at the same settings. The supernatant was removed by aspiration and cell pellets were resuspended in 1 mL RPMI-C and pipetted through 70 µM filters (PluriSelect #43-10070-70). Filtered PBMCs proceeded immediately to staining for the unstimulated panel or to overnight stimulation for the AIM panel.

### Cell Surface Staining for Detection of Activation Induced Markers (AIM)

Activation induced cell marker (AIM) expression was assessed by flow cytometry as previously described(50). All timepoints and experimental conditions ((PHA positive control, DMSO negative controls, stimulation with non-structural (NON) peptide pool, stimulation with structural (STR) peptide pool)) for each subject were included in the same experimental run.

Filtered PBMCs were seeded 50 µL per well in 96-well U-bottom plates followed by the addition of 4X treatments. Each clinical sample was stimulated with the following treatments: RPMI-C only control (“UNS”), phytohemagglutinin (“PHA”, Roche #11249738001, 10.0 µg/mL) as a positive control, polio structural megapool (“STR”,1.0 µg/mL), polio non-structural megapool (“NON”, 1.0 µg/mL), and triplicate wells of dimethyl sulfoxide (“DMSO”, Sigma #D8418) equimolar to the peptide megapools as a negative vehicle control. In addition to the polio megapools, a SARS-CoV-2 spike megapool (“COV”, 1.0 µg/mL) was used to stimulate the donor control. The donor control samples were PBMCs from a single blood draw on one individual that were aliquoted into many cryovials, and one cryovial was included in each round of stimulation, staining, and sample acquisition. To stimulate cells for single-color controls, PBMCs were plated from a single individual with the same treatments described above. Using cells from a single individual allowed cells to be pooled following stimulation and then divided to ensure a mixture of activation states in each single-color control to capture the brightest protein expression for all markers. Plates were incubated 24 hours at 37 degrees Celsius and 5% CO2. To generate a sample specific unstained control for autofluorescence extraction (“unstained”), as well as the a4β7 AF700 fluorescence minus one (“FMO”) control, 20 µL of cells were removed from the various treatment wells of a single sample, pooled, and plated into two wells for the respective staining protocols. Staining was performed in 96-well U-bottom plates for samples, donor control, FMO, and unstained controls and in in 1.5 mL Eppendorf tubes for single-color controls.

Following overnight stimulation, plates were centrifuged and supernatants discarded. Samples, donor control, and FMO were resuspended in 50 µL of UV Blue LiveDead (Invitrogen #L34962, 1:200), unstained was resuspended in 50 µL of phosphate buffered saline (“PBS”, Corning #21-031-CV), and incubated at 4 degrees Celsius. To wash, 100 µL PBS 1% FBS was added to each well and plates were centrifuged and supernatants removed. The antibody cocktail was made in Brilliant Buffer (BD Horizon #566349) containing blocking antibodies (Fc Block, BioLegend #422302, 1:20; Monocyte Block, BioLegend #426103, 1:20) and all fluorescent antibodies (CD69 BUV661, BD #750213, 1:100; CD8 BUV805, BD #612890, 1:400; CD4 Pacific Blue, BioLegend #317424, 1:400; CD3 BV510, BioLegend #344828, 1:50; CD134/OX40 BV605, BioLegend #350028, 1:20; CD14 BV711, BioLegend #301838, 1:100; CD19 BV711, BioLegend #302246, 1:100; CXCR5 BV750, BioLegend #356942, 1:20; CD279/PD-1 PE, BioLegend #329906, 1:40; CD45 PerCP/Cy5.5, BioLegend #368504, 1:50; CD137 PE/Cy7, BioLegend #309818, 1:50; TCRgd APC, BioLegend #331212, 1:20) except a4β7AF700. Once 100 µL was set aside as the FMO cocktail, the final antibody was added (a4β7 AF700, R+D #FAB10078N, 1:40). Samples and donor control were resuspended in antibody cocktail, FMO in FMO cocktail, and unstained in PBS, plates were incubated at 4 degrees Celsius. Samples were washed in PBS 1% FBS, centrifuged and supernatants removed. For fixation, samples, donor control, FMO, and unstained were resuspended in PBS 1% paraformaldehyde (Alfa Aesar #43386) and incubated at 4 degrees Celsius. A final wash of PBS 1% FBS was added to each well, plates were centrifuged, supernatants removed, samples resuspended in PBS, plates wrapped tightly with parafilm, and stored at 4 degrees Celsius. Immediately before acquisition, PBS was added to each well and samples were thoroughly mixed to resuspend cells.

The single-color control stains were prepared in 1.5 mL Eppendorf tubes at the dilutions indicated above, using PBS 1% FBS for all antibodies and the negative control, and PBS for UV Blue LIVE/DEAD. Following overnight stimulation, cells stimulated for the single-color controls were pooled in a reservoir. An aliquot of cells was removed from the reservoir into a tube and heated at 65 degrees Celsius for 10 minutes. An equal aliquot of cells was added to the tube of heat-killed cells, and the tube was centrifuged, supernatant aspirated and resuspended in the UV Blue LIVE/DEAD stain. The remaining cells in the reservoir were replated, centrifuged, and supernatants removed. Using 50 µL of the prepared stains, cell pellets were resuspended in the plate wells and transferred to tubes, incubated at 4 degrees Celsius. Samples were washed in PBS 1% FBS, centrifuged, and supernatant aspirated. Cell pellets were resuspended in 1% paraformaldehyde and incubated at 4 degrees Celsius. A final wash of PBS 1% FBS was added to each tube and centrifuged, and supernatant removed. Cells were resuspended in 400 µL PBS and transferred to capped 5 mL round bottom tubes for storage at 4 degrees Celsius.

### Flow Cytometry Data Acquisition

Samples were acquired on the Aurora Cytek under settings set by passing daily QC, with the following adjustments: forward scatter 60, side scatter 220, threshold 300,000, forward area scaling 1.0. Between 100,000-200,000 events were collected into the lymphocyte scatter gate for all single-color controls. To account for changes in autofluorescence over days of storage, the sample specific unstained control was acquired (25,000 events) each day that samples were acquired and applied to the live unmixing for those samples (only for AIM Panel). For the Unstimulated Panel, samples were run in tubes at a flow rate ∼2,000 events/second, with a stop gate set to 500,000 events in the lymphocyte scatter gate. For the AIM Panel, samples were run in plates at a flow rate ∼5-10,000 events/second, with stop gates set for time (2 minutes), volume (300 µL), and count (500,000 events into the lymphocyte scatter gate). Similar to the sample specific unstained control, the a4β7 AF700 FMO was acquired (25,000 events) each day that samples were acquired (only for AIM Panel).

### Flow Cytometry Gating

The online cytometry analysis platform OMIQ (Dotmatics) was used for manual bidimensional gating of raw flow cytometry data. FCS files were uploaded to OMIQ into individual workflows corresponding to each experimental run. Scaling and gating were performed uniformly across all experimental runs. Scaling was first adjusted on all fluorescent features by using an Arcsinh transformation. A time gate was then individually adjusted for each FCS file, followed by application of a universally applied hierarchical gating scheme. PHA stimulated samples were scaled and gated separately. Additional scaling and gating for AIM+ (CD137+OX40+) CD4+ T cells and AIM+ (CD137+CD69+) CD8+ T cell gates was done at the plate level to account for experimental variation in fluorescence signal detection (see **Fig S1A**, gating scheme). Data, including cell counts and percent (%) parent gate, were then exported as .csv files and read into R studio for downstream analysis (code available at https://github.com/P-Harvey/Ag_Spec_T_cell_PV).

### Antigen-specific T cell Analysis

Following initial assay development in which the expression of various AIM makers were assessed after stimulation protocols in our laboratory, we down selected to those AIM markers with consistent and robust expression readily detectible in combination with our complete fluorophore antibody panel. For CD4+ T cells, CD137+OX40+ was chosen, and for CD8+ T cells CD137+CD69+ was chosen. The frequency of poliovirus-specific AIM+ (CD137+OX40+CD4+) and (CD137+CD69+CD8+) T cells was separately determined for each peptide stimulation condition (structural and non-structural peptide pools) using a background subtraction (BS) method, as previously reported(20). Briefly, the frequency (%) of AIM+ T cells was calculated by subtracting the percent (%) AIM+ cells detected in sample-specific DMSO negative vehicle controls (run in triplicate and averaged) from the percent (%) of AIM+ cells detected for each sample and AIM+ parameter combination assessed. If the averaged DMSO value was < LOS, the parameter-specific LOS value was used.

The assay limit of detection (LOD) was calculated for each AIM parameter by taking twice the upper bound of the 95% CI of the geometric mean of the arithmetic means 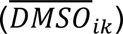 of DMSO triplicates and taking twice the upper bound of the 95% CI. Any averaged DMSO values < 0.005 were imputed to 0.005. The LOD was calculated to be 0.0166% for AIM+CD4+ and 0.022% for AIM+CD8+ parameters.

The assay limit of sensitivity (LOS) was calculated for each AIM parameter as two standard deviations above the median of the averaged DMSO triplicates. The LOS was calculated to be 0.021% for AIM+CD4+ and 0.033% for AIM+CD8+ parameters.

The stimulation index (SI) was calculated for each sample and T cell/AIM+ parameter combination as the frequency (%) of AIM+ cells divided by the averaged DMSO values per parameter condition.

Samples with a BS value > LOD and > LOS, and with a SI > 2 met response criteria and were considered to represent a positive T cell response. AIM+ T cell frequencies for samples that did not meet positive response criteria were subsequently set to “0”.

Total poliovirus-specific CD4+ and CD8+ T cells were calculated for samples that met response criteria by summing the frequency (%) of background subtracted AIM+ cells detected in structural and non-structural stimulation conditions per sample.

### Calculation of Absolute Cell Counts

To estimate the absolute counts of Ag. specific T cells in whole blood, we first multiplied the absolute Lymphocyte count (provided by the UVMMC clinical laboratory for the corresponding paired peripheral blood sample (obtained at the same blood draw) by the Pct. (%) CD4+ (or CD8+) of Lymphocytes (as determined by flow cytometry). This value was then multiplied by the background subtracted percentage of parent (% CD4/CD137+OX40+ of CD4+ or CD8/CD137+CD69+ of CD8+) for each sample and treatment condition (stimulation with non-structural or structural peptide pools), giving a treatment specific estimate of Ag. specific T cells per sample (cells/mcL). If a LAN responded to both treatments (non-structural and structural), the mean of the two treatments’ cell counts were used in calculating the values in Table S6; if a sample responded to only one treatment, then those exact cell counts were used. Negative treatment responses were not used in the estimate of absolute cell counts.

### Statistical Analysis

Unless otherwise noted, all statistical comparisons used two-tailed tests to determine whether there was any difference between two measures. In general, comparisons of mean values were conducted using Welch’s two sample t test, and comparisons of proportions were conducted using an exact binomial test.

For Welch’s two sample t test we satisfied the assumption that the data are distributed approximately normally, testing the null hypothesis that the mean of group A was equal to group B with the alternative that the means were different between the two groups.

For the exact Binomial test we satisfied the assumptions that the data are randomly sampled, the samples were independent, and the outcome measure was binary (success vs. failure). We tested the null hypothesis that the probability of success in group A was equal to the probability of success in group B, with the alternative that the probability of success differed between the two groups.

In certain comparisons (Fig. 3C,F), the Clopper-Pearson method was used to measure the uncertainty in our estimation of proportions. The specific implementation for this method was taken from the PropCIs (v0.3-1) R package using the exactci function.

We controlled for multiple comparisons by calculating q-values using the Storey-Tibshirani procedure to account for dependency between comparisons. We controlled the False Discovery Rate (pFDR) at ≤ 10%. The q-value represents the probability that a given comparison is significant, given the total number of statistical comparisons made in these exploratory analyses.

Descriptive statistics were used to calculate proportions, frequencies, and mean values of variables of interest. Correlations were made using R^2^ values to measure the amount of residual error relative to the error explained by the regression ((SSR – SSE) / SSR). Confidence Intervals were calculated as the mean plus or minus the critical value times the quotient of standard deviation and the square root of the number of samples (standard error). In general, the critical value was from a student t distribution on n – 1 degrees of freedom and α = 0.05. Geometric means were calculated using the DescTools R package.

All statistical analyses were conducted in RStudio Version 2024.09.0 Build 375 (using R version 4.4.1). The source code for these analyses and additional documentation are available at https://github.com/P-Harvey/Poliovirus_Antigen_Specific_TCells.

## Reagent List

Histopaque-1077 (Sigma #H8889)

Cell Freezing Media (Sigma #C6164)

RPMI 1640 (Cytiva #SH30096.02)

Heat-Inactivated Fetal Bovine Serum (Corning #35-011-CV)

L-Glutamine (Gibco #A2916802)

Penicillin-Streptomycin (Gibco #15070063)

Benzonase (Sigma-Aldrich #E1014-25KU)

70 µM Filters (PluriSelect #43-10070-70)

Phytohemagglutinin PHA (Roche #11249738001)

Dimethyl Sulfoxide (“DMSO”, Sigma #D8418)

Phosphate Buffered Saline (“PBS”, Corning #21-031-CV)

1% Paraformaldehyde (Alfa Aesar #43386)

UV Blue LiveDead (Invitrogen #L34962, 1:200)

Brilliant Buffer (BD Horizon #566349)

Fc Block (BioLegend #422302, 1:20)

Monocyte Block (BioLegend #426103, 1:20)

CD69 BUV661 (BD #750213, 1:100)

CD8 BUV805 (BD #612890, 1:400)

CD4 Pacific Blue (BioLegend #317424, 1:400)

CD3 BV510 (BioLegend #344828, 1:50)

CD134/OX40 BV605 (BioLegend #350028, 1:20)

CD14 BV711 (BioLegend #301838, 1:100)

CD19 BV711 (BioLegend #302246, 1:100)

CXCR5 BV750 (BioLegend #356942, 1:20)

CD279/PD-1 PE (BioLegend #329906, 1:40)

CD45 PerCP/Cy5.5 (BioLegend #368504, 1:50)

CD137 PE/Cy7 (BioLegend #309818, 1:50)

TCRgd APC (BioLegend #331212, 1:20)

a4β7 AF700 (R+D #FAB10078N, 1:40)

CD45RA BUV395 (BD OptiBuild #740315, 1:200)

CD20 BUV563 (BD OptiBuild #748456, 1:800)

CD196/CCR6 BUV661 (BD OptiBuild #750696, 1:400)

CD8 BUV805 (BD Horizon #612890, 1:100)

CD197/CCR7 BV421 (BioLegend #353208, 1:40)

CD4 PacBlue (BioLegend #317423, 1:400)

CD19 BV480 (BD #568214, 1:100)

CD3 BV510 (BioLegend #344828, 1:100)

IgM BV570 (BioLegend #314517, 1:100)

CD14 BV711 (BioLegend #301838, 1:100)

CD185/CXCR5 BV750 (BioLegend #356942, 1:100)

IgA VioBright B515 (MACS Miltenyi #130-116-886, 1:200)

MR1 5-OP-RU PE (Emory/NIH #NA, 1:100)

CD27 PE-CF594 (BD #562297, 1:100)

CD45 PerCP/Cy5.5 (BioLegend #368504, 1:40)

CD183/CXCR3 PE/Cy7 (BioLegend #353720, 1:100)

TCRgd APC (BioLegend #331211, 1:20)

a4β7 AF700 (R+D #FAB10078N, 1:40)

CD38 APC/Fire810 (BioLegend #303549, 1:100)

