## Supplemental Figures for "Oral live attenuated polio vaccines induce enhanced T-cell responses with broad antigen recognition compared to inactivated polio vaccines"

### Figure S1: Overlap of structural and non-structural epitope reactivity per sample

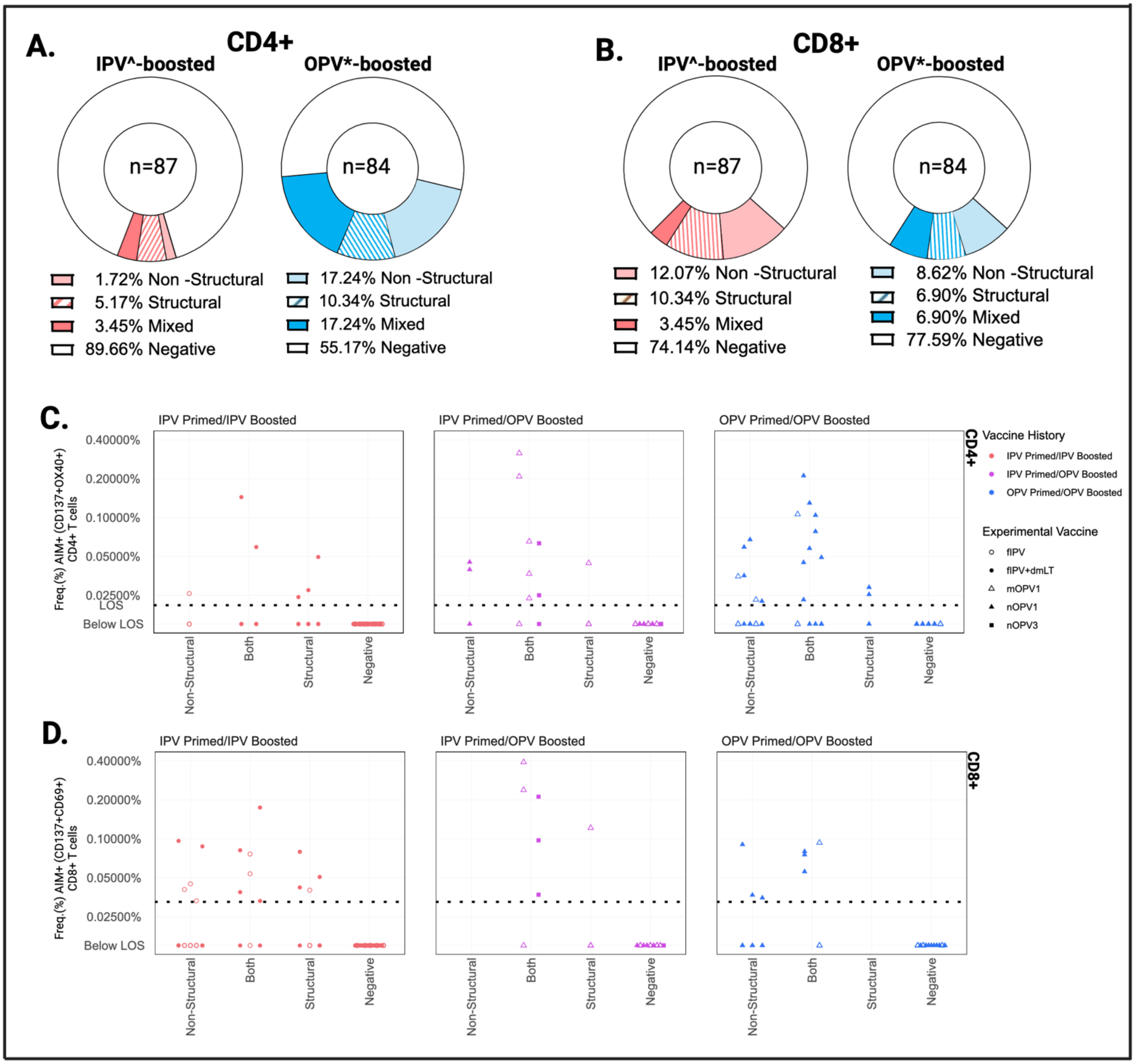


**Figure S1**: **A.** The proportion of samples (n) with AIM+CD4+ **(A)** and/or CD8+ **(B)** T cells that met response criteria following stimulation with non-structural peptide pools alone (lighter solid), structural peptide pools alone (striped), or both (mixed, darker solid). Frequency of total poliovirus-specific CD4+ (**C**) and CD8+ **(D)** T cells detected per sample. Values (%) are background subtracted and summed for those samples that reacted to both peptide pools ("mixed"). **E.** Bivariate distribution of each sample that met response criteria depicted according to CD4+/CD8+ response on x-axis and nonstructural/structural response on y-axis (min/max scaling). Each relative proportion was given a value between 0 and 1. Dots are colored by vaccine history and sized by sample time point. NON, nonstructural; STR, structural; LOD, limit of detection. IPV^ formulations include tIPV with or without dmLT; OPV* formulations include nOPV1, mOPV1, nOPV3, mOPV3

### Figure S2: Correlation of CD4+ & CD8+ T cell responses with shedding index endpoint

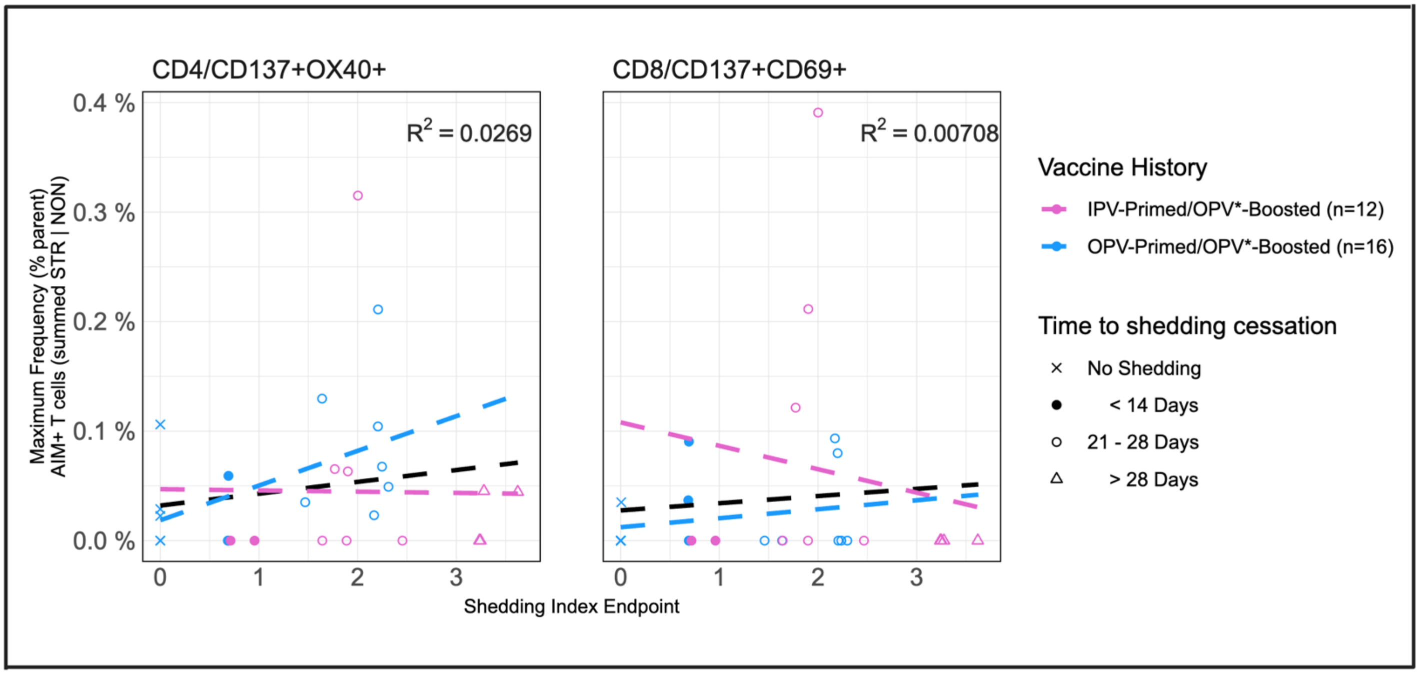


**Figure S2**: Correlation of shedding index endpoint and maximum frequency of total AIM+ (CD137+OX40+) CD4+ and (CD137+CD69+) CD8+ T cells detected per subject. Values (%) are background subtracted and summed (STR, structural; NON, nonstructural). Regression lines provided for all samples (black; with $R^2$ value) and for each prime/boost group. Data points are shaped according to time to shedding cessation and colors according to vaccine history (prime/boost). OPV* formulations include nOPV1, mOPV1, nOPV3, mOPV3

### Figure S3: CD4+ & CD8+ T cell responses according to fecal viral shedding duration

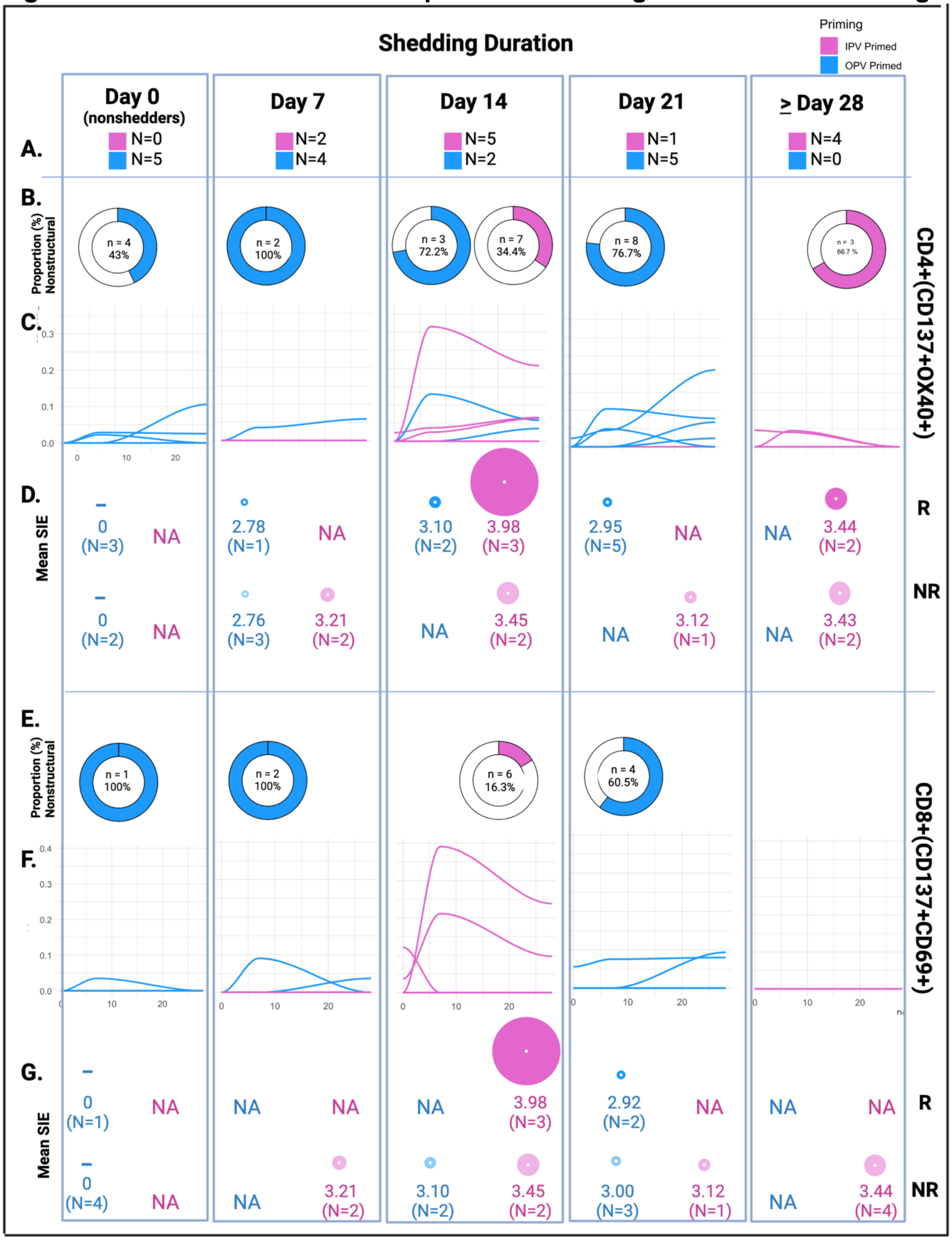


**Figure S3. A.** IPV and OPV primed volunteers (N) were categorized according to the time point of their last positive stool sample. **B, E.** Doughnuts represent the proportion of the total CD4+ (B) and CD8+ (D) T cell response directed towards non-structural viral proteins across all samples (n) meeting response criteria from volunteers within each respective grouping. **C, F**. The longitudinal CD4+ (C ) and CD8+ (E ) antigen-specific T cell responses for each volunteer. **D, G** Dots are scaled to represent the relative mean SIE value of volunteers with (AIM +) T cell responses (R, Responder) and without (NR, Non-responder) within each group. Number of volunteers included in each group is represented (N). Data is colored according to polio vaccine priming history. SIE, shedding index endpoint; AIM, antigen-induced markers; NA, not applicable (denotes no volunteers included in grouping)
