## Supplemental Tables for "Oral live attenuated polio vaccines induce enhanced T-cell responses with broad antigen recognition compared to inactivated polio vaccines"

Supplementary Tables

**Table S1:** Demographics of volunteers enrolled in polio vaccine studies

| **Study** | **IPV-boosted Study ( NCT04529538)** | | | **OPV-boosted Study (NCT03922061)** | | | | **Combined Total** |
|---|---|---|---|---|---|---|---|---|
| **Boosting (Vaccine type)** | **fIPV only** | **fIPV + dmLT** | **Total** | **mOPV1** | **nOPV1** | **nOPV3** | **Total** | |
| **Sample Size** n/N (%) | | | | | | | | |
| Volunteers | 10/29 (35.5%) | 19/29 (65.5%) | 29/57 (50.9%) | 10/28 (35.7%) | 16/28 (57.1%) | 2/28 (7.1%) | 28/57 (49.1%) | 57/57 (100.0%) |
| **Priming History** n/N (%) | | | | | | | | |
| IPV | 10/10 (100.0%) | 19/19 (100.0%) | 29/29 (100.0%) | 6/10 (60.0%) | 4/16 (25.0%) | 2/ 2 (100.0%) | 12/28 (42.9%) | 41/57 (72.0%) |
| OPV | – | – | – | 4/10 (40.0%) | 12/16 (75.0%) | – | 16/28 (57.1%) | 16/57 (28.0%) |
| **Ethnicity** n/N (%) | | | | | | | | |
| Non-Hispanic | 9/10 (90.0%) | 17/19 (89.5%) | 26/29 (89.7%) | 10/10 (100.0%) | 15/16 (93.8%) | 2/ 2 (100.0%) | 27/28 (96.4%) | 53/57 (93.0%) |
| Hispanic | 1/10 (10.0%) | 2/19 (10.5%) | 3/29 (10.3%) | 0/10 (0.0%) | 1/16 (6.3%) | 0/ 2 (0.0%) | 1/28 (3.6%) | 4/57 (7.0%) |
| **Race** n/N (%) | | | | | | | | |
| White | 9/10 (90.0%) | 19/19 (100.0%) | 28/29 (96.6.%) | 9/10 (90.0%) | 16/16 (100.0%) | 2/ 2 (100.0%) | 27/28 (96.4%) | 55/57 (96.5%) |
| Black/African American | 1/10 (10.0%) | 0/19 (0.0%) | 1/29 (3.4%) | 1/10 (10.0%) | 0/16 (0.0%) | 0/ 2 (0.0%) | 1/28 (3.6%) | 2/57 (3.5%) |
| **Sex** n/N (%) | | | | | | | | |
| Male | 1/10 (10.0%) | 7/19 (36.8%) | 8/29 (27.6%) | 5/10 (50.0%) | 7/16 (43.8%) | 1/ 2 (50.0%) | 13/28 (46.4%) | 21/57 (36.8%) |
| Female | 9/10 (90.0%) | 12/19 (63.2%) | 21/29 (72.4%) | 5/10 (50.0%) | 9/16 (56.3%) | 1/ 2 (50.0%) | 15/28 (53.6%) | 36/57 (63.2%) |
| **Age** Years | | | | | | | | |
| Mean (SD) | 18.8 (0.632) | 19.0 (0.667) | 18.9 (0.651) | 25.2 (7.52) | 28.0 (5.56) | 20 (1.41) | 26.4 (6.42) | 22.6 (5.86) |
| Median | 19 | 19 | 19 | 22.5 | 27.5 | 20 | 25.5 | 20 |
| Min, Max | 18, 20 | 18, 20 | 18, 20 | 19, 40 | 20, 39 | 19, 21 | 19, 40 | 18, 40 |

IPV: inactivated poliovirus vaccine; OPV: oral poliovirus vaccine; fIPV: fractional dose trivalent inactivated poliovirus vaccine IPV; dmLT: LT(R192G/L211A; n: sample number; N: total samples; mOPV1: monovalent Sabin strain oral poliovirus vaccine serotype 1; nOPV1: monovalent novel oral poliovirus vaccine serotype 1; nOPV3: monovalent novel oral poliovirus vaccine serotype 3; SD: standard deviation; min: minimum; max: maximum.

Supplementary Tables

**Table S2: Serotype specific serum neutralizing antibody (SNA) titers**

| Study | IPV-boosted Study (NCT04529538) | | | OPV-boosted Study (NCT03922061) | | | | Combined Total |
|---|---|---|---|---|---|---|---|---|
| | **fIPV only** | **fIPV + dmLT** | **Total** | **mOPV1** | **nOPV1** | **nOPV3** | **Total** | |
| **Serotype-specific SNA titers** Geometric Mean Titer ($log_2$) (min. Titer, max. Titer) | | | | | | | | |
| **PV1** | | | | | | | | |
| Baseline | 5.657 (2.50, 9.17) | 4.446 (2.50, 10.50) | 4.831 (2.50, 10.50) | 10.248 (8.50, 10.50) | 5.893 (2.50, 9.83) | 8.333 ( 8.17, 8.50) | 6.003 (2.50, 9.83) | 5.364 (2.50, 10.50) |
| Day 28 | 9.739 (7.50, 10.50) | 10.037 (6.83, 10.50) | 9.934 (6.83, 10.50) | 10.248 (8.50, 10.50) | 10.306 (9.50, 10.50) | 8.138 ( 7.50, 8.83) | 10.113 (7.50, 10.50) | 10.026 (6.83, 10.50) |
| **PV2** | | | | | | | | |
| Baseline | 5.719 (3.17, 10.50) | 5.502 (3.17, 10.50) | 5.576 (3.17, 10.50) | 6.746 (2.83, 10.17) | 6.126 (2.50, 10.50) | 5.347 ( 3.50, 8.17) | 6.263 (2.50, 10.50) | 5.897 (2.50, 10.50) |
| Day 28 | 10.043 (9.17, 10.50) | 10.072 (8.17, 10.50) | 10.062 (8.17, 10.50) | 8.721 (7.17, 10.50) | 7.993 (2.50, 10.50) | 5.191 ( 3.17, 8.50) | 7.995 (2.50, 10.50) | 8.970 (2.50, 10.50) |
| **PV3** | | | | | | | | |
| Baseline | 6.979 (2.50, 10.50) | 6.856 (2.50, 10.50) | 6.898 (2.50, 10.50) | 6.313 (2.50, 10.50) | 5.074 (2.50, 10.50) | 7.576 ( 6.50, 8.83) | 5.622 (2.50, 10.50) | 6.250 (2.50, 10.50) |
| Day 28 | 10.343 (9.17, 10.50) | 10.338 (9.50, 10.50) | 10.340 (9.17, 10.50) | 7.505 (2.50, 10.50) | 6.218 (2.50, 10.50) | 10.500 (10.50, 10.50) | 6.904 (2.50, 10.50) | 8.449 (2.50, 10.50) |

IPV: inactivated poliovirus vaccine; OPV: oral poliovirus vaccine; fIPV: fractional dose trivalent inactivated poliovirus vaccine IPV; dmLT: LT(R192G/L211A; mOPV1: monovalent Sabin strain oral poliovirus vaccine serotype 1; nOPV1: monovalent novel oral poliovirus vaccine serotype 1; nOPV3: monovalent novel oral poliovirus vaccine serotype 3; PV1: poliovirus serotype 1; PV2: poliovirus serotype 2; PV3: poliovirus serotype 3; min: minimum; max: maximum.

### Table S3: CD4+ and CD8+ T cell responses by day

| **Boosting (Study) (Vaccine type)** | **IPV-boosted Study (NCT04529538) (fIPV +/- dmLT)** | | | **OPV[a] -boosted Study (NCT03922061)** | | | **Total (N = 57)** | **$Comp_1$** | **$Comp_2$** |
|---|---|---|---|---|---|---|---|---|---|
| **Priming History** | **IPV (N = 29)** | **OPV (N = 0)** | **IPV Study All (N = 29)** | **IPV (N = 12)** | **OPV (N = 16)** | **OPV Study All (N = 28)** | | **p-value q-value** | **p-value q-value** |
| **Number (n/N (%)) of volunteers with CD4+ T cell response by timepoint** | | | | | | | | | |
| Baseline | 3 (10.3%) | – | 3 (10.3%) | 2 (16.7%) | 1 ( 6.3%) | 3 (10.7%) | 6 (10.5%) | 1.0000 1.0000 | 1.0000 1.0000 |
| Day 7 | 0 ( 0.0%) | – | 0 ( 0.0%) | 5 (41.7%) | 7 (43.8%) | 12 (42.9%) | 12 (21.1%) | 0.7744 0.9152 | **0.0005** |
| Day 10 | 3 (10.3%) | – | 3 (10.3%) | – | – | – | 3 (10.3%)* | – | – |
| Day 28 | – | – | – | 3 (25.0%) | 9 (56.3%) | 12 (42.9%) | 12 (42.9%) | 0.1460 0.4195 | – |
| All | 6 (20.7%) | – | 6 (20.7%) | 5 (41.7%) | 11 (68.8%) | 16 (57.1%) | 22 (38.6%) | 0.2101 0.4419 | 0.0525 0.2275 |
| **Mean[b] (min, max) frequency (% parent) of CD4+ T cell response by timepoint** | | | | | | | | | |
| Baseline | 0.073 (0.026, 0.144) | – | 0.073 (0.026, 0.144) | 0.034 (0.024, 0.045) | 0.023 (0.023, 0.023) | 0.031 (0.023, 0.045) | 0.052 (0.023, 0.144) | – | 0.3607 0.6116 |
| Day 7 | 0.000 (0.000, 0.000) | – | 0.000 (0.000, 0.000) | 0.092 (0.025, 0.315) | 0.059 (0.023, 0.123) | 0.073 (0.023, 0.315) | 0.073 (0.022, 0.315) | 0.5966 0.7506 | 0.0161 0.0897 |
| Day 10 | 0.037 (0.024, 0.059) | – | 0.037 (0.024, 0.059) | – | – | – | 0.037 (0.024, 0.059) | – | – |
| Day 28 | – | – | – | 0.112 (0.063,0.208) | 0.074 (0.023, 0.211) | 0.083 (0.023, 0.211) | 0.083 (0.023, 0.211) | 0.5157 0.6968 | – |
| All | 0.055 (0.024, 0.144) | – | 0.055 (0.024, 0.144) | 0.087 (0.024, 0.315) | 0.065 (0.023, 0.211) | 0.073 (0.023, 0.315) | 0.070 (0.023, 0.315) | 0.5181 0.6968 | 0.4555 0.6707 |
| **Number (n/N (%)) of volunteers with CD8+ T cell response by timepoint** | | | | | | | | | |
| Baseline | 8 (27.6%) | – | 8 (27.6%) | 2 (16.7%) | 1 ( 6.3%) | 3 (10.7%) | 11 (19.3%) | 1.0000 1.0000 | 0.2266 0.4419 |
| Day 7 | 4 (13.8%) | – | 4 (13.8%) | 2 (16.7%) | 3 (18.8%) | 5 (17.9%) | 9 (15.8%) | 1.0000 1.0000 | 0.2668 0.4730 |
| Day 10 | 3 (10.3%) | – | 3 (10.3%) | – | – | – | 3 (10.3%)* | – | – |
| Day 28 | – | – | – | 2 (16.7%) | 3 (18.8%) | 5 (17.9%) | 5 (17.9%)* | 1.0000 1.0000 | – |
| All | 11 (37.9%) | – | 11 (37.9%) | 3 (25.0%) | 5 (31.3%) | 8 (28.6%) | 19 (33.3%) | 0.7266 0.8855 | 0.2005 0.4419 |
| **Mean[b] (min, max) frequency of CD8+ T cell response by timepoint** | | | | | | | | | |
| Baseline | 0.071 (0.039, 0.174) | – | 0.071 (0.039, 0.174) | 0.079 (0.037, 0.121) | 0.056 (0.056, 0.056) | 0.071 (0.037, 0.121) | 0.071 (0.037, 0.174) | – | 0.9799 1.0000 |
| Day 7 | 0.068 (0.033, 0.082) | – | 0.068 (0.033, 0.082) | 0.301 (0.211, 0.391) | 0.067 (0.035, 0.090) | 0.161 (0.035, 0.391) | 0.119 (0.033, 0.391) | 0.2234 0.4419 | 0.2255 0.4419 |
| Day 10 | 0.046 (0.033, 0.054) | – | 0.046 (0.033, 0.054) | – | – | – | 0.046 (0.033, 0.054) | – | – |
| Day 28 | – | – | – | 0.168 (0.097, 0.238) | 0.070 (0.037, 0.093) | 0.109 (0.037, 0.238) | 0.109 (0.037, 0.238) | – | – |
| All | 0.065 (0.033, 0.174) | – | 0.065 (0.033, 0.174) | 0.183 (0.037, 0.391) | 0.067 (0.035, 0.093) | 0.120 (0.035, 0.391) | 0.091 (0.033, 0.391) | 0.0740 0.2886 | 0.0856 0.3035 |

IPV: inactivated poliovirus vaccine; OPV: oral poliovirus vaccine; fIPV: fractional dose trivalent inactivated poliovirus vaccine IPV; dmLT: LT(R192G/L211A;n: sample number; N: total samples; min: minimum; max: maximum.

[a] Inclusive of all OPV vaccine variations (monovalent and novel; PV1 and PV3)

[b] Means and proportions inclusive only of samples with CD4+ reactivity detected

[c] Frequency = summed background subtracted pct. of parent only for samples meeting response criteria

"_" denotes no samples collected: No day 28 for Study #1, no OPV Primed in Study #1, No day 10 for Study #2

$Comp_1$ is comparison of IPV vs OPV Priming within the OPV-boosted study

$Comp_2$ is comparison of IPV vs OPV Boosting across studies

Supplementary Tables

**Table S4: Proportion of samples with CD4+ and CD8+ T cells reactive to structural, non-structural, or both peptide megapools**

| | Baseline | Day 7 | Day 10 | Day 28 | Total |
|---|---|---|---|---|---|
| **Number (n / N (%))** of samples with CD4+ T cells reactive to Structural peptide megapool alone (Prime/Boost) | | | | | |
| IPV/IPV | 1/29 ( 3.4%) | 0/29 ( 0.0%) | 2/29 ( 6.9%) | _ | 3/ 87 ( 3.5%) |
| IPV/OPV | 1/12 ( 8.3%) | 2/12 (16.7%) | _ | 1/12 ( 8.3%) | 4/ 36 (11.1%) |
| OPV/OPV | 0/16 ( 0.0%) | 1/16 ( 6.2%) | _ | 1/16 ( 6.2%) | 2/ 48 ( 4.2%) |
| All | 2/57 ( 3.5%) | 3/57 ( 5.3%) | 2/29 ( 6.9%) | 2/28 ( 7.1%) | 9/171 ( 5.3%) |
| **Number (n / N (%))** of samples with CD4+ T cells reactive to Nonstructural peptide megapool alone | | | | | |
| IPV/IPV | 1/29 ( 3.4%) | 0/29 ( 0.0%) | 0/29 ( 0.0%) | _ | 1/ 87 ( 1.2%) |
| IPV/OPV | 1/12 ( 8.3%) | 2/12 (16.7%) | _ | 0/12 ( 0.0%) | 3/ 36 ( 8.3%) |
| OPV/OPV | 1/16 ( 6.2%) | 3/16 (18.8%) | _ | 4/16 (25.0%) | 8/ 48 (16.7%) |
| All | 3/57 ( 5.3%) | 5/57 ( 8.8%) | 0/29 ( 0.0%) | 4/28 (14.3%) | 12/171 ( 7.0%) |
| **Number (n / N (%))** of samples with CD4+ T cells reactive to both Structural and Non-structural peptide megapools | | | | | |
| IPV/IPV | 1/29 ( 3.4%) | 0/29 ( 0.0%) | 1/29 ( 3.4%) | _ | 2/ 87 ( 2.3%) |
| IPV/OPV | 0/12 ( 0.0%) | 1/12 ( 8.3%) | _ | 2/12 (16.7%) | 3/ 36 ( 8.3%) |
| OPV/OPV | 0/16 ( 0.0%) | 3/16 (18.8%) | _ | 4/16 (25.0%) | 7/ 48 ( 9.0%) |
| All | 1/57 ( 1.8%) | 4/57 ( 7.0%) | 1/29 ( 3.4%) | 6/28 (21.4%) | 12/171 ( 7.0%) |
| **Total** | 6/57 (10.5%) | 12/57 (21.1%) | 3/29 (10.3%) | 12/28 (42.9%) | 33/171 (19.3%) |
| **Number (n / N (%))** of samples with CD8+ T cells reactive to Structural peptide megapool alone | | | | | |
| IPV/IPV | 2/29 ( 6.9%) | 1/29 ( 3.4%) | 3/29 (10.3%) | _ | 6/ 87 ( 6.9%) |
| IPV/OPV | 2/12 (16.7%) | 1/12 ( 8.3%) | _ | 0/12 ( 0.0%) | 3/ 36 ( 8.3%) |
| OPV/OPV | 1/16 ( 6.2%) | 0/16 ( 0.0%) | _ | 0/16 ( 0.0%) | 1/ 48 ( 2.1%) |
| All | 5/57 ( 8.8%) | 2/57 ( 3.5%) | 3/29 (10.3%) | 0/28 ( 0.0%) | 10/171 ( 5.9%) |
| **Number (n / N (%))** of samples with CD8+ T cells reactive to Nonstructural peptide megapool alone | | | | | |
| IPV/IPV | 6/29 (20.7%) | 1/29 ( 3.4%) | 0/29 ( 0.0%) | _ | 7/ 87 ( 8.1%) |
| IPV/OPV | 0/12 ( 0.0%) | 0/12 ( 0.0%) | _ | 0/12 ( 0.0%) | 0/ 36 ( 0.0%) |
| OPV/OPV | 0/16 ( 0.0%) | 3/16 (18.8%) | _ | 2/16 (12.5%) | 5/ 48 (10.4%) |
| All | 6/57 (10.5%) | 4/57 ( 7.0%) | 0/29 ( 0.0%) | 2/28 ( 7.1%) | 12/171 ( 7.0%) |
| **Number (n / N (%))** of samples with CD8+ T cells reactive to both Structural and Non-structural peptide megapools | | | | | |
| IPV/IPV | 0/29 ( 0.0%) | 2/29 ( 6.9%) | 0/29 ( 0.0%) | _ | 2/ 87 ( 2.3%) |
| IPV/OPV | 0/12 ( 0.0%) | 1/12 ( 8.3%) | _ | 2/12 (16.7%) | 3/ 36 ( 8.3%) |
| OPV/OPV | 0/16 ( 0.0%) | 0/16 ( 0.0%) | _ | 1/16 ( 6.2%) | 1/ 48 ( 2.1%) |
| All | 0/57 ( 0.0%) | 3/57 ( 5.3%) | 0/29 ( 0.0%) | 3/28 (10.7%) | 6/171 ( 3.5%) |
| **Total** | 11/57 (19.3%) | 9/57 (15.8%) | 3/29 (10.3%) | 5/28 (17.9%) | 28/171 (16.4%) |

IPV: inactivated poliovirus vaccine; OPV: oral poliovirus vaccine; n: sample number; N: total samples.

Supplementary Tables

**Table S5: Mean frequency of antigen-specific CD4+ and CD8+ T cells detected in samples reactive to structural, non-structural, or both peptide megapools**

| | | | | | |
|---|---|---|---|---|---|
| **Mean (Min, Max)** frequency of ag-specific CD4+ T cells (% of CD4+ T cells) detected in samples reactive to Structural peptide megapool alone (n = 9 / 33) | | | | | |
| IPV/IPV | 0.0495 (0.0495, 0.0495) | – | 0.0258 (0.0242, 0.0274) | – | 0.0337 (0.0242, 0.0495) |
| IPV/OPV | 0.0237 (0.0237, 0.0237) | 0.0406 (0.0367, 0.0444) | – | 0.0632 (0.0632, 0.0632) | 0.0420 (0.0237, 0.0632) |
| OPV/OPV | – | 0.0288 (0.0288, 0.0288) | – | 0.0254 (0.0254, 0.0254) | 0.0271 (0.0254, 0.0288) |
| All | 0.0366 (0.0237, 0.0495) | 0.0367 (0.0288, 0.0444) | 0.0258 (0.0242, 0.0274) | 0.0443 (0.0254, 0.0632) | 0.0360 (0.0237, 0.0632) |
| **Mean (Min, Max)** frequency of ag-specific CD4+ T cells (% of CD4+ T cells) detected in samples reactive to Non-structural peptide megapools alone (n = 12 / 33) | | | | | |
| IPV/IPV | 0.0258 (0.0258, 0.0258) | – | – | – | 0.0258 (0.0258, 0.0258) |
| IPV/OPV | 0.0452 (0.0452, 0.0452) | 0.0323 (0.0250, 0.0396) | – | – | 0.0366 (0.0250, 0.0452 |
| OPV/OPV | 0.0231 (0.0231, 0.0231) | 0.0343 (0.0225, 0.0449) | – | 0.0462 (0.0231, 0.0675) | 0.0389 (0.0225, 0.0675) |
| All | 0.0314 (0.0231, 0.0452) | 0.0335 (0.0225, 0.0449) | – | 0.0462 (0.0231, 0.0675) | 0.0372 (0.0225, 0.0675) |
| **Mean (Min, Max)** frequency of ag-specific CD4+ T cells (% of CD4+ T cells) detected in samples reactive to both Structural and Non-structural peptide megapools [1] (n = 12 / 33) | | | | | |
| IPV/IPV | 0.1441 (0.1441, 0.1441) | – | 0.0582 (0.0592, 0.0592) | – | 0.1016 (0.0592, 0.1441) |
| IPV/OPV | – | 0.3151 (0.3151, 0.3151) | – | 0.1367 (0.0654, 0.2080) | 0.1962 (0.0654, 0.3151) |
| OPV/OPV | – | 0.0944 (0.0492, 0.1300) | – | 0.1132 (0.0578, 0.2111) | 0.1051 (0.0492, 0.2111) |
| All | 0.1441 (0.1441, 0.1441) | 0.1495 (0.0492, 0.3150) | 0.0582 (0.0592, 0.0592) | 0.1211 (0.0578, 0.2111) | 0.1273 (0.0492, 0.3151) |
| **Mean proportion (%)**[2] of total CD4+ T cell response reactive to Non-structural peptides in samples with both Structural and Non-structural responses detected (n = 12 / 33) | | | | | |
| IPV/IPV | 70.83% | _ | 35.55% | _ | 53.19% |
| IPV/OPV | _ | 67.30% | _ | 36.86% | 47.01% |
| OPV/OPV | _ | 58.24% | _ | 56.87% | 57.46% |
| All | 70.83% | 60.50% | 35.55% | 50.20% | 54.13% |
| **Mean (Min, Max)** frequency of ag-specific CD8+ T cells (% of CD8+ T cells) detected in samples reactive to Structural peptide megapool alone (n = 10 / 28) | | | | | |
| IPV/IPV | 0.0411 (0.0401, 0.0421) | 0.0794 (0.0794, 0.0794) | 0.0459 (0.0331, 0.0537) | – | 0.0499 (0.0331, 0.0794) |
| IPV/OPV | 0.0792 (0.0370, 0.1210) | 0.2114 (0.2114, 0.2114) | – | – | 0.1233 (0.0370, 0.2114) |
| OPV/OPV | 0.0558 (0.0558, 0.0558) | – | – | – | 0.0558 (0.0558, 0.0558) |
| All | 0.0593 (0.0370, 0.1210) | 0.1454 (0.0794, 0.2114) | 0.0459 (0.0331, 0.0537) | – | 0.0725 (0.0331, 0.2114) |
| **Mean (Min, Max)** frequency of ag-specific CD8+ T cells (% of CD8+ T cells) detected in samples reactive to Nonstructural peptide megapool alone (n = 12 / 28) | | | | | |
| IPV/IPV | 0.0804 (0.0387, 0.1740) | 0.0332 (0.0332, 0.0332) | – | – | 0.0737 (0.0332, 0.1744) |
| IPV/OPV | – | – | – | – | – |
| OPV/OPV | – | 0.0670 (0.0350, 0.0904) | – | 0.0583 (0.0368, 0.0799) | 0.0635 (0.0350, 0.0904) |
| All | 0.0804 (0.0387, 0.1740) | 0.0585 (0.0332, 0.0904) | – | 0.0583 (0.0368, 0.0799) | 0.0694 (0.0332, 0.1740) |
| **Mean (Min, Max)** frequency of ag-specific CD8+ T cells (% of CD8+ T cells) detected in samples reactive to both Structural and Non-structural peptide megapools[1] (n = 6 / 28) | | | | | |
| IPV/IPV | – | 0.0789 (0.0763, 0.0815) | – | – | 0.0789 (0.0763, 0.0815) |

Supplementary Tables

| | | | | | |
|---|---|---|---|---|---|
| IPV/OPV | _ | 0.3908 (0.3908, 0.3908) | _ | 0.1678 (0.0973, 0.2380) | 0.2422 (0.0973, 0.3908) |
| OPV/OPV | _ | _ | _ | 0.0933 (0.0933, 0.0933) | 0.0933 (0.0933, 0.0933) |
| All | _ | 0.1829 (0.0763, 0.3908) | _ | 0.1430 (0.0933, 0.2380) | 0.1630 (0.0763, 0.3908) |
| **Mean proportion (%)**[2] of total CD8+ T cell response reactive to Non-structural peptides in samples with both Structural and Non-structural responses detected | | | | | |
| IPV/IPV | _ | 53.05% | _ | _ | 53.05% |
| IPV/OPV | _ | 32.86% | _ | 36.62% | 32.70% |
| OPV/OPV | _ | _ | _ | 42.13% | 42.13% |
| All | _ | 46.32% | _ | 35.79% | 41.05% |

IPV: inactivated poliovirus vaccine; OPV: oral poliovirus vaccine; ag: antigen; n: sample number; min: minimum; max: maximum.

[1] Summed Back. Sub. Mean (min., max.) is used for samples responding to both Structural and Non-Structural
[2] Mean(Non-Structural / Summed Back. Sub.) for samples responding to both Structural and Non-Structural
"_" denotes no samples collected: No day 28 for Study #1, no OPV Primed in Study #1, No day 10 for Study #2

Supplementary Tables

### Table S6: Absolute cell counts calculated for samples with CD4+ and CD8+ T cell responses

| OPV-boosted Study (NCT03922061) | | | | | | |
|---|---|---|---|---|---|---|
| **Priming/Boosting** | **Mean Abs. Lymph. per mcL** | **Mean CD4+ T-Cell Pct. Lymph.** | **Mean Abs. CD4+ T-Cells per mcL** | **Mean CD4+ Ag(+) T cells Pct. Parent** | **Mean Abs. CD4+ Ag(+) T cells per mcL** | **Expected Abs. CD4+ Ag(+) T cells per 8 mL** |
| **Day 0** | | | | | | |
| IPV Primed/OPV Boosted (n = 2 /12) | 2265 | 44.37% (37.245, 51.485) | 990 (920, 1061) | 0.03% (0.024, 0.045) | 0.34 (0.20, 0.52) | 2693 (1600, 4160) |
| OPV Primed/OPV Boosted (n = 1/16) | 1550 | 35.09% (35.094, 35.094) | 544 (544, 544) | 0.02% (0.023, 0.023) | 0.13 (0.13, 0.13) | 1001 (1001, 1001) |
| **All (n = 3/28)** | 2027 | 41.28% (35.094, 51.485) | 841 (544, 1061) | 0.03% (0.023, 0.045) | 0.26 (0.16, 0.47) | 2086 (1280, 3760) |
| **Day 7** | | | | | | |
| IPV Primed/OPV Boosted (n = 5/12) | 2132 | 48.84% (34.529, 57.702) | 1059 (656, 1575) | 0.09% (0.025, 0.315) | 0.97 (0.18, 3.88) | 7794 (1440, 31040) |
| OPV Primed/OPV Boosted (n = 7/16) | 1736 | 47.53% (36.345, 63.267) | 808 (428, 1097) | 0.06% (0.023, 0.13) | 0.48 (0.15, 1.43) | 3814 (1200, 11440) |
| **All (n = 12/28)** | 1901 | 48.08% (34.529, 63.267) | 913 (428, 1575) | 0.07% (0.023, 0.315) | 0.67 (0.15, 3.79) | 5332 (1200, 30320) |
| **Day 28** | | | | | | |
| IPV Primed/OPV Boosted (n = 3/12) | 2077 | 52.49% (49.492, 54.469) | 1094 (728, 1585) | 0.11% (0.063, 0.208) | 1.23 (0.65, 2.35) | 9802 (5200, 18800) |
| OPV Primed/OPV Boosted (n = 9/16) | 1624 | 45.60% (35.514, 65.884) | 737 (504, 1021) | 0.07% (0.023, 0.211) | 0.55 (0.13, 2.26) | 4363 (1040, 18080) |
| **All (n = 12/28)** | 1738 | 47.33% (35.514, 65.884) | 826 (504, 1585) | 0.08% (0.023, 0.211) | 0.69 (0.14, 2.42) | 5485 (1120, 19360) |
| **All time-points** | | | | | | |
| IPV Primed/OPV Boosted (n = 10/36) | 2142 | 49.04% (34.529, 57.702) | 1056 (656, 1585) | 0.09% (0.024, 0.315) | 0.92 (0.18, 3.89) | 7350 (1440, 31120) |
| OPV Primed/OPV Boosted (n = 17/48) | 1666 | 45.78% (35.094, 65.884) | 755 (428, 1097) | 0.07% (0.023, 0.211) | 0.49 (0.13, 2.32) | 3926 (1040, 18560) |
| **All (n = 27/84)** | 1842 | 46.99% (34.529, 65.884) | 866 (428, 1585) | 0.07% (0.023, 0.315) | 0.63 (0.15, 3.82) | 5057 (1200, 30560) |
| **Priming/Boosting** | **Mean Abs. Lymph. per mcL** | **Mean CD8+ T-Cell Pct. Lymph.** | **Mean Abs. CD8+ T-Cells per mcL** | **Mean CD8+ Ag(+) T cells Pct. Parent** | **Mean Abs. CD8+ Ag(+) T cells per mcL** | **Expected Abs. CD8+ Ag(+) T cells per 8 mL** |
| **Day 0** | | | | | | |
| IPV Primed/OPV Boosted (n = 2/12) | 1875 | 15.25% (13.911, 16.593) | 283 (280, 287) | 0.08% (0.037, 0.121) | 0.22 (0.10, 0.38) | 1789 (800, 3040) |
| OPV Primed/OPV Boosted (n = 1/16) | 1550 | 21.76% (21.762, 21.762) | 337 (337, 337) | 0.06% (0.056, 0.056) | 0.19 (0.19, 0.19) | 1510 (1510, 1510) |
| **All (n = 3/28)** | 1767 | 17.42% (13.911, 21.762) | 301 (280, 337) | 0.07% (0.037, 0.121) | 0.21 (0.09, 0.47) | 1710 (720, 3760) |
| **Day 7** | | | | | | |
| IPV Primed/OPV Boosted (n = 2/12) | 1895 | 15.93% (15.56, 16.297) | 302 (294, 310) | 0.30% (0.211, 0.391) | 0.91 (0.62, 1.21) | 7272 (4960, 9680) |
| OPV Primed/OPV Boosted (n = 3/16) | 1543 | 20.03% (16.318, 22.503) | 320 (162, 468) | 0.07% (0.035, 0.09) | 0.21 (0.09, 0.31) | 1715 (720, 2480) |
| **All (n = 5/28)** | 1684 | 18.39% (15.56, 22.503) | 313 (162, 468) | 0.16% (0.035, 0.391) | 0.50 (0.09, 1.48) | 4031 (720, 11840) |
| **Day 28** | | | | | | |
| IPV Primed/OPV Boosted (n = 2/12) | 1660 | 17.43% (15.632, 19.223) | 284 (261, 306) | 0.17% (0.097, 0.238) | 0.48 (0.25, 0.76) | 3817 (2000, 6080) |
| OPV Primed/OPV Boosted (n = 3/16) | 1607 | 18.08% (14.794, 20.481) | 292 (225, 358) | 0.07% (0.037, 0.093) | 0.20 (0.09, 0.31) | 1635 (720, 2480) |
| **All (n = 5/28)** | 1628 | 17.82% (14.794, 20.481) | 289 (225, 358) | 0.11% (0.037, 0.238) | 0.32 (0.09, 0.79) | 2520 (720, 6320) |
| **All time-points** | | | | | | |
| IPV Primed/OPV Boosted (n = 6/36) | 1810 | 16.20% (13.911, 19.223) | 290 (261, 310) | 0.18% (0.037, 0.391) | 0.53 (0.09, 1.36) | 4246 (720, 10880) |
| OPV Primed/OPV Boosted (n = 7/48) | 1571 | 19.44% (14.794, 22.503) | 311 (162, 468) | 0.07% (0.035, 0.093) | 0.21 (0.08, 0.33) | 1667 (640, 2640) |
| **All (n = 13/84)** | 1682 | 17.95% (13.911, 22.503) | 301 (162, 468) | 0.12% (0.035, 0.391) | 0.36 (0.08, 1.48) | 2890 (640, 11840) |

### Supplementary Tables

mcL: microliter; Abs: absolute; lymph: lymphocyte; pct: percent; ag: antigen; IPV: inactivated poliovirus vaccine; OPV: oral poliovirus vaccine; n: sample number.

Calculations only include samples that met response criteria; N will therefore be different at each timepoint

Supplementary Tables

**Table S7: Proportion of volunteers with fecal poliovirus shedding detected by day**

| OPV-boosted Study (NCT03922061) | | | | |
|---|---|---|---|---|
| **Priming History** | **Proportion (n/N) study participants with PCR-positive fecal sample** | | | |
| | **Day 7** | **Day 14** | **Day 21** | **Day 28** |
| IPV-primed | 12/12 (100.0 %) | 10/12 (83.3%) | 5/12 (41.7%) | 4/12 (33.3%) |
| OPV-primed | 11/16 ( 68.8%) | 7/16 (43.8%) | 5/16 (31.3%) | 0/16 ( 0.0%) |
| All | 23/28 ( 82.1%) | 17/28 (60.7%) | 10/28 (35.7%) | 4/28 (14.3%) |
| | **Mean $\log_{10}$ $CCID_{50}$ viral titer** | | | |
| | **Day 7** | **Day 14** | **Day 21** | **Day 28** |
| IPV-primed | 4.24 (3.83, 4.69) | 2.92 (2.71, 3.15) | 2.85 (2.57, 3.17) | 2.95 (2.38, 3.67) |
| OPV-primed | 3.15 (2.9, 3.41) | 2.77 (2.72, 2.83) | 2.75 (2.75, 2.75) | 0.00 (0.00, 0.00) |
| All | 3.67 (3.36, 4.02) | 2.86 (2.74, 2.99) | 2.80 (2.69, 2.92) | 2.95 (2.38, 3.67) |

IPV: inactivated poliovirus vaccine; OPV: oral poliovirus vaccine; n: sample number; N: total samples

[a] Inclusive of all volunteers enrolled in Study #2 and all OPV vaccine variations (monovalent and novel; PV1 and PV3).

[b] The titer value provided matches the vaccine serotype (poliovirus type 1 or 3) received by the volunteer. We considered any poliovirus type 1 or 3-specific stool neutralization titer >2 to be detectable.

Supplementary Tables

### Table S8: Time to Shedding Cessation and Shedding Index Endpoint by priming cohort

| OPV-boosted Study (NCT03922061) | | | | |
|---|---|---|---|---|
| **Boosting (Vaccine type)** | **OPV[a]** | | | **Comp$_5$** |
| **Priming History** | **IPV (n = 12)** | **OPV (n = 16)** | **All (n = 28)** | **p-value q-value** |
| **Time to shedding cessation[b] (Days)** | | | | |
| Mean (95% CI) | 26.833 (20.040, 33.627) | 14.875 ( 8.652, 21.098) | 20.000 (15.116, 24.884) | **3.475x10$^{-11}$ 3.823x10$^{-10}$** |
| min | 14 | 0 | 0 | |
| max | 42 | 28 | 42 | |
| **Shedding Index Endpoint[c] (SIE)** | | | | |
| Mean (95% CI) | 2.229 ( 1.627, 2.831) | 1.063 ( 0.557, 1.568) | 1.563 ( 1.136, 1.989) | **0.0004 0.0039** |
| min | 0.719 | 0.000 | 0.000 | |
| max | 2.305 | 3.625 | 3.625 | |

IPV: inactivated poliovirus vaccine; OPV: oral poliovirus vaccine; n: sample number; SIE: shedding index endpoint; CI: confidence interval; min: minimum; max: maximum.

[a] Inclusive of all OPV vaccine variations (monovalent and novel; PV1 and PV3).

[b] Time to shedding cessation was defined as the collection date of the first stool samples that tested negative for the presence of poliovirus by PCR. Shedding cessation was confirmed by two stool samples collected at least 24 hours apart that tested negative by PCR.

[c] SIE is calculated using $log_{10}$ $CCID_{50}$ titer results from shared stool collection timepoints (Days 7, 14, 21, 28)

**Comp$_5$** is comparison of IPV vs OPV Priming within Study #2

**Comp$_5$** is calculated using Welch two-sample t test for difference of means.

**Table S9: Time to Shedding Cessation and Shedding Index Endpoint by T cell response**

| **OPV [d] -boosted Study (NCT03922061)** | | | | | | |
|---|---|---|---|---|---|---|
| **Boosting (Vaccine type)** | **OPV [d]** | | | | **Comp$_6$** p-value q-value | **Comp$_7$** p-value q-value |
| **Priming History** | **IPV** | | **OPV** | | | |
| **T cell response[a]** | **(n = 5) (+)** | **(n = 7) (–)** | **(n = 12) (+)** | **(n = 4) (–)** | | |
| **Time to shedding cessation [b] (Days)** | | | | | | |
| Mean (95% CI) | 29.40 (15.118, 43.682) | 25.00 (15.212, 34.788) | 17.50 (10.034, 24.966) | 7 (-5.862, 19.862) | 0.0772 0.1545 | **<0.00005 <0.00005** |
| min | 21 | 14 | 0 | 0 | | |
| max | 42 | 42 | 21 | 14 | | |
| **Shedding Index Endpoint (SIE)[c]** | | | | | | |
| Mean (95% CI) | 2.52 (1.44, 3.59) | 2.02 (1.09, 2.96) | 1.30 (0.69, 1.91) | 0.35 (-0.29, 0.98) | 0.3895 0.6329 | **0.0156 0.0897** |
| min | 1.77 | 0.72 | 0.00 | 0.00 | | |
| max | 3.63 | 3.25 | 2.30 | 0.70 | | |

IPV: inactivated poliovirus vaccine; OPV: oral poliovirus vaccine; n: sample number; SIE: shedding index endpoint; CI: confidence interval; min: minimum; max: maximum.

[a] T cell response is inclusive of detection of CD4+ and/or CD8+ T cell reactivity at any timepoint that met response criteria

[b] Time to shedding cessation was defined as the collection date of the first stool samples that tested negative for the presence of poliovirus by PCR. Shedding cessation was confirmed by two stool samples collected at least 24 hours apart that tested negative by PCR.

[c] SIE is calculated using $log_{10}$ $CCID_{50}$ titer results from shared stool collection timepoints (Days 7, 14, 21, 28)

[d] Inclusive of all OPV vaccine variations (monovalent and novel; PV1 and PV3).

**Comp$_6$** is comparison of T cell (+) vs T cell (–) within IPV Primed cohort

**Comp$_7$** is comparison of T cell (+) vs T cell (–) within OPV Primed cohort

Comparisons calculated using Welch two-sample t test for difference of means.
